# Gene-exposure interactions regulate cytokine-mediated chronic inflammation and cardiac remodeling

**DOI:** 10.64898/2026.01.19.26344405

**Authors:** Mattia Corianò, Shamin Tahasildar, Ling Huang, Khaled Rjoob, Soodeh Kalaie, Jin Zheng, Lara Curran, Parisa Gifani, Marc-Emmanuel Dumas, Declan P O’Regan

## Abstract

**Background:** Chronic inflammation predicts adverse cardiovascular outcomes, but mechanisms linking systemic inflammation to cardiac remodeling remain incompletely understood. We investigated associations between circulating inflammatory biomarkers and cardiac phenotypes in a population-based cohort and examined how environmental exposures and genetic susceptibility influence inflammatory responses.

**Methods:** We analyzed subsets of 488,079 UK Biobank participants with metabolomic and proteomic profiling, cardiac magnetic resonance (CMR) imaging, and longitudinal outcomes. Chronic inflammation was quantified using glycoprotein acetyls (GlycA) by nuclear magnetic resonance spectroscopy. Machine learning-based analysis extracted CMR phenotypes. Multivariable linear regression assessed GlycA-cardiac associations. Mediation analysis tested 80 inflammatory proteins as potential mediators. Cox models evaluated GlycA levels and major adverse cardiovascular events (MACE). An exposome-wide association study identified environmental determinants of inflammation, and gene-environment interactions were assessed using multi-ancestry polygenic risk scores.

**Results:** Higher GlycA levels were associated with restrictive cardiac remodeling: reduced left ventricular indexed end-diastolic volume (β = –2.09) and stroke volume (β = –1.12) with compensatory increased heart rate (β = 1.38; all *P* < 10^−228^). Interleukin (IL) −1 receptor antagonist mediated 27% of the GlycA effect on end-diastolic volume (average causal mediated effect –0.51 [95% CI, –0.53 to –0.64]; *P* < 10^−16^). The highest GlycA quintile had 43% higher MACE risk versus the lowest (adjusted HR, 1.43 [95% CI, 1.38–1.49]). Trunk fat mass (β = 0.35), current smoking (β = 0.39), psychological distress, and low socioeconomic status were the strongest GlycA determinants (all *P* < 10^−50^). Cardiovascular polygenic risk scores modified associations between environmental exposures, inflammation, and MACE.

**Conclusions:** Chronic systemic inflammation is associated with restrictive cardiac remodeling and increased cardiovascular risk mediated by circulating cytokines and growth factors. Individual inflammatory responses are shaped by gene-environment interactions, highlighting the complex interplay between genetic susceptibility, environmental exposures, and their cumulative impact on cardiovascular health.

## Introduction

Chronic low-grade inflammation has emerged as a clinically actionable mediator of cardiovascular disease (CVD). Residual inflammatory risk, independent of traditional lipid-based factors, predicts adverse outcomes and represents a therapeutic target for both primary and secondary prevention.^1^ There is also growing evidence that systemic inflammation is a hallmark of non-ischemic heart failure (HF), particularly when complicated by cardio-metabolic disease.^2,3^ However, the interacting mechanisms triggering, regulating and responding to systemic inflammation in the adult population have not been addressed by previous biomarker outcome studies.^4–6^ Consequently, neither inflammation-associated cardiac phenotypes nor the specific proteins mediating cardiac adaptation have been systematically characterized at population scale.

Equally important are the environmental and genetic determinants of chronic inflammation. While individual environmental exposures have been linked to inflammation and mortality,^7–9^ we lack comprehensive exposome-wide analyses that identify key environmental drivers and their interaction with genetic susceptibility.^10^ Given that combined assessments of clinical, metabolomic, and polygenic scores offer improved cardiovascular risk prediction,^11^ discovering the causes, consequences, and modifiers of systemic inflammation is needed to support individualized prevention and the prioritization of new therapeutic strategies in diverse risk groups.

Taking advantage of large-scale assays of stable inflammatory biomarkers in deeply-phenotyped populations with multi-omic profiling, we sought to determine the structural, proteomic, and environmental underpinnings of inflammatory risk. Using circulating glycoprotein acetyls (GlycA) as a stable and sensitive composite marker of chronic inflammation,^12^ we characterized the predominant pattern of cardiac remodeling through machine learning analysis of cardiovascular imaging. To establish a comprehensive risk framework, we assessed the prognostic link between GlycA and major adverse cardiovascular events (MACE) in the community, resolved the specific protein mediators of this association, and mapped the interaction between the exposome and multi-ancestry polygenic risk scores (PRS) in driving systemic inflammation. By defining the environmental, genetic, and phenotypic architecture of chronic inflammation, we aim to provide the evidence base necessary for precision cardiovascular risk stratification and therapeutic prioritization in risk-diverse populations.

## Methods

### Study overview

The UK Biobank (UKB) study enrolled approximately 500,000 community-dwelling participants aged 40–69 years from across the United Kingdom, between 2006 and 2010.^13^ All participants provided written informed consent, and the study received approval from the National Research Ethics Service (11/NW/0382). Our analysis was conducted under access approval number 40616, with data last accessed on December 1, 2025.

An overview of the methods is shown in Figure 1. We used nuclear magnetic resonance (NMR) metabolomics data in 488,079 participants to quantify circulating GlycA.^14^ Within this group, we assessed the relationship between GlycA and incident outcomes. To investigate the relationship between systemic inflammation and cardiac remodeling, we employed machine learning techniques to derive 33 imaging-derived phenotypes (IDP)s of cardiac structure and function from cardiac magnetic resonance (CMR) and 16 electrocardiogram (ECG) traits in a subset of 70,809 participants.^15^ This analysis enabled us to determine associations between GlycA levels and quantitative measures of cardiac morphology and function.

**Figure 1.**
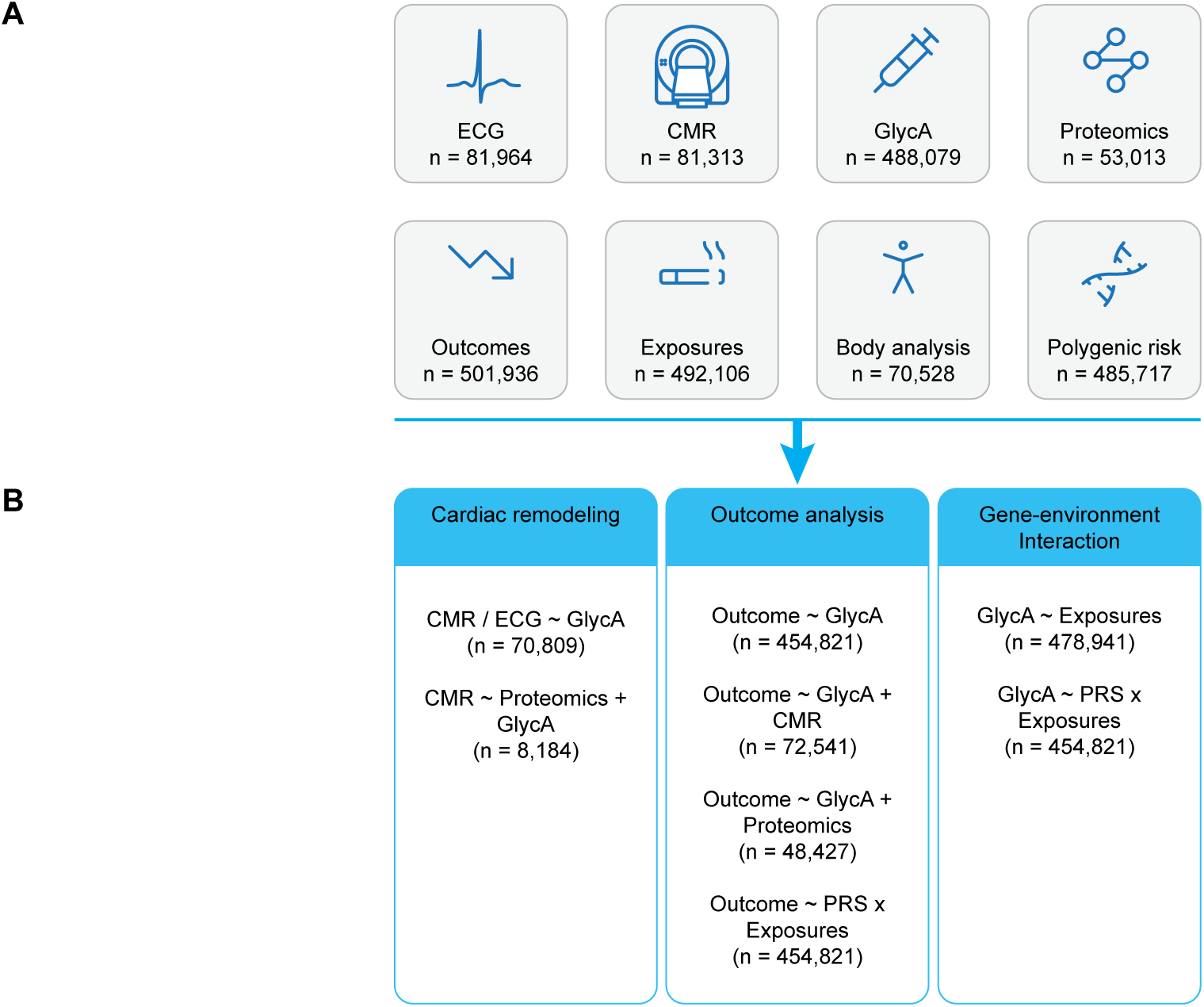
Study flowchart. **a)** The study included UK Biobank participants with metabolomic and proteomic profiling, CMR imaging, ECG, environmental exposures, body fat analysis, polygenic risk profiles and longitudinal outcomes. Chronic inflammation was quantified using GlycA by nuclear magnetic resonance spectroscopy. **b)** The relationship between systemic inflammation and cardiac remodeling was assessed in a subset of 70,809 participants considering 33 IDPs from CMR and 16 ECG traits. Mediation analysis tested 80 inflammatory proteins as potential mediators in 8,184 individuals. Outcome analysis investigated the association of GlycA on MACE and all-cause death on top of lipid profile (454,821), IDPs (72,541), and considering proteins as mediators (48,427). An exposome-wide association study on 478,941 volunteers identified environmental determinants of inflammation, and gene-environment interactions were assessed using multi-ancestry polygenic risk scores available for 454,821 participants. **Abbreviations:** CMR, cardiac magnetic resonance; ECG, electrocardiogram; GlycA, glycoprotein acetyls; IDP, imaging-derived phenotype; MACE, major adverse cardiac events; PRS, polygenic risk score.

We further investigated the proteomic landscape underlying GlycA elevation by analyzing 80 proteins, selected a priori based on their established roles in systemic inflammatory processes.^16^ These proteins were assessed in 52,503 participants with available metabolomics and proteomics profiles. Each protein was evaluated both as a predictor of GlycA levels and as a potential mediator of associations between GlycA and cardiac IDPs or clinical outcomes. Mediation analysis was used to determine the proportion of GlycA’s effect that was explained through each protein pathway.

To identify environmental determinants of systemic inflammation, we performed an exposome-wide association study (XWAS) in 478,941 participants, examining 169 environmental exposures and 8 body composition measurements in relation to GlycA levels.^7^ Exposures demonstrating significant associations with inflammation were subsequently analyzed for interactions with 10 multi-ancestry cardiovascular disease PRSs. Gene-environment interactions were tested using interaction terms in multivariable linear regression models for GlycA levels and Cox proportional hazards models for MACE.

### Blood biomarkers

UKB provides measurements of a comprehensive range of biochemical markers from biological samples collected at baseline (2006-2010) across all 500,000 participants. For this study, we analyzed variables from the biochemistry, metabolite, and proteomic biomarker panels.

Biochemistry biomarkers were obtained from 473,133 participants at baseline recruitment. Technical details of quantitative analyses have been previously described.^17^ We analyzed the following biochemistry biomarkers: creatinine (field ID 30700), HbA1c (field ID 30750), High-sensitivity C-reactive protein (hsCRP) (field ID 30710), and lipoprotein (a) (Lp(a)) (field ID 30790).

NMR metabolomics measures were obtained from 488,079 participants recruited at baseline and profiled by Nightingale Health. Technical details of the quantitative serum NMR analysis have been previously described.^18^ Pre-processing included removal of technical variation, re-calculation of derived biomarker, removal of outlier values, normal distribution transformation and standardization as described in Supplementary Methods. For this study, we considered GlycA (field ID 23480) as marker of chronic inflammation. Data from the November 2025 release was used in this study.

Proteomic profiling was obtained from 53,013 participants recruited at baseline and performed using the Olink Explore 3072 platform, which integrates four Olink panels (Cardiometabolic, Inflammation, Neurology, and Oncology). UKB Olink data are provided as normalized protein expression values on a log2 scale. For the present study, we focused on 80 biomarkers from the Olink Inflammation panel (Supplementary Table 1), previously shown to be associated with adverse cardiac structure and function in smaller observational studies.^16^

### Cardiovascular imaging analysis

CMR imaging was performed on a 1.5T scanner (MAGNETOM Aera, Siemens Healthineers, Erlangen, Germany) using retrospectively gated balanced steady-state free precession cine sequences.^19^ Left ventricular short-axis cine images were acquired from base to apex, along with long-axis cine images in the two-chamber and four-chamber planes. Cines comprised 50 cardiac phases with a typical temporal resolution of 31 ms. Transverse cine images of the ascending and descending thoracic aorta were also acquired.

Images were segmented using fully convolutional neural networks, with segmentation quality equivalent to expert human readers.^20^ Derived measurements of the left and right ventricles were determined from the two-dimensional images: indexed end-diastsolic volume (EDVi), indexed end-systsolic volume (ESVi), stroke volume, and ejection fraction (EF). Left ventricular mass was calculated from myocardial volume using an assumed myocardial density of 1.05 g/mL. Atrial volumes were derived using the biplane area-length method: 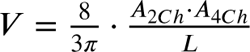, where *A_2Ch_* and *A_4Ch_* represent the atrial areas on the two-chamber and four-chamber views, respectively, and *L* is the averaged longitudinal diameter across the two views. CMR-derived measurements were indexed to body surface area (BSA) calculated using the Du Bois formula: BSA = 0.007184 × Height^0.725^ × Weight^0.425^, with height in cm and weight in kg. Left ventricular wall thickness was measured as the distance between the segmented epicardium and endocardium at end-diastole.

Myocardial strain analysis was performed using non-rigid free-form deformation image registration between successive frames.^21^ To reduce accumulation of registration errors, motion tracking was performed in both forward and backward directions from the end-diastolic frame, and an average displacement field was calculated.^22^ Circumferential (*E*_*cc*_) and radial (*E_rr_*) strains were calculated using short-axis cines as 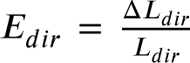, where *dir* represents the circumferential or radial direction, *L*_*dir*_ is the absolute length of a line segment along this direction, and Δ*L*_*dir*_ is its change in length over time. Longitudinal (*E*_*ll*_) strain was calculated from long-axis four-chamber motion tracking. Strain rate was estimated as the first derivative of strain, and peak-diastolic strain rate (PDSR) in the radial and longitudinal directions was detected using an algorithm to identify local maxima.^22^

A spatio-temporal neural network was used to segment the aortic cine images.^15^ Maximum and minimum cross-sectional areas were derived, and aortic distensibility was calculated using central blood pressure obtained from peripheral pulse-wave analysis (Vicorder, SMT Medical, Würzburg, Germany).^23^ A total of 33 IDPs characterizing cardiac volumes and function, as well as vascular stiffness were extracted.

### Electrocardiogram

Twelve-lead ECG recordings were obtained from UKB participants using a standardized acquisition protocol and analyzed with proprietary software (GE CardioSoft). Raw 15-second, 12-lead ECG signals from 81,964 participants were processed through a validated computational pipeline to systematically extract quantitative ECG biomarkers. This analysis yielded 16 distinct ECG parameters for subsequent cardiovascular risk assessment.

### Environmental factors

We assessed the impact of environmental exposures on systemic inflammation using the analytical framework developed by Argentieri et al.^7^ From 176 baseline exposure variables available as of August 1, 2025, we retained variables with <45% missing data that were consistently collected across all assessment centers. This yielded 148 candidate exposures for analysis.

Prior to analysis, all continuous variables were centered and standardized, with the exception of age at recruitment. Ordinal categorical variables were recoded to enable linear association testing, while nominal categorical exposures were analyzed using the most frequent category as the reference group. Multi-select (“mark all that apply”) responses were converted to binary dummy variables to facilitate statistical modeling. The final analytical dataset exhibited an average missing data rate of 6% across all variables (range: 0–44%).

We addressed missing values using the R package missRanger, which employs random forest imputation combined with predictive mean matching. Imputation parameters included a maximum of 10 iterations, 200 trees per random forest, and 16 processing threads, with remaining hyperparameters using default settings. Following imputation and variable recoding, 169 environmental exposures were available for XWAS analysis.

Comprehensive data dictionaries detailing all XWAS exposures are provided in Supplementary Table 2, with variable recoding procedures described in the Supplementary Methods.

### Body composition

Abdominal and whole-body adipose tissue assessment was performed using 1.5T dual-echo Dixon Vibe imaging from neck to knees. Six overlapping sections underwent calibration, stacking, fusion, and segmentation using AMRA Researcher software (AMRA Medical AB, Linköping, Sweden). Android and gynoid adipose tissue masses were quantified using self-supervised, multi-modal alignment between magnetic resonance imaging and dual-energy X-ray absorptiometry scans.^24^ Liver proton density fat fraction was measured using the least median of squares Dixon method during single breath-hold acquisitions.^25^ For this study, we considered 8 body fat composition measurements reflecting total fat and specific adiposity patterns as shown in Supplementary Table 3.

### Exposome wide association study

XWAS analyses were performed using multivariable linear regression with log-transformed and scaled GlycA as the outcome variable, regressed against 177 factors (169 environmental exposures + 8 body composition measurements). Each model was stratified by age at recruitment (5-year birth cohorts) and sex, and adjusted for BSA, sex, age, age*²*, age:sex interaction, and ethnicity (White, Asian, Black, or Other). All continuous exposure variables were scaled and centered prior to analysis.

For each model, β coefficients were calculated separately within each stratum, with resulting effect estimates representing those that provided the best fit across all strata. Analyses were conducted for the entire cohort (pooled cohort) and independently in each sex. Parallel XWAS analyses were performed using hsCRP as the outcome variable instead of GlycA.

*P* values were corrected for multiple testing using the false discovery rate (FDR) method (Benjamini–Hochberg procedure^26^) with a significance threshold of FDR *P* < 0.05.

### Outcome data

UKB provides comprehensive health outcome information from multiple linked data sources. In this study, health outcomes were ascertained from self-reported medical history (field IDs 20002, 20008), inpatient hospital records (field IDs 41270 and 41280 for ICD-10 codes; 41271 and 41281 for ICD-9 codes; 20010 for operation codes), first occurrence datasets, and national death registries. The primary endpoint was the first occurrence of MACE, defined as a composite of cardiac arrest, HF, stroke or other cerebrovascular events, myocardial infarction, or cardiovascular death (Supplementary Table 4). All-cause mortality was investigated as a secondary endpoint. Follow-up commenced at enrollment and continued until the first occurrence of MACE or death for metabolomics analyses, or from CMR acquisition until MACE or death for imaging-based analyses, depending on the specific research question.

### Gene-environment interactions

We investigated gene-environment interactions using multi-ancestry PRSs for all-cause CVD, serum lipid levels, diabetes, hypertension and stroke (Supplementary Table 5). Details regarding the derivation of these PRS have been previously described.^27^ We examined whether interactions between environmental factors and each PRS influence GlycA levels using multivariable linear regression models, and incident MACE using Cox proportional hazards models, as described in the Supplementary Methods. A secondary sensitivity analyses further adjusted for Townsend deprivation index at recruitment (field ID 22189) was performed.

### Statistical analysis

Detailed statistical methods are provided in the Supplementary Methods. All analyses were performed using R version 4.5.0 and Python version 3.9.

Continuous variables are presented as mean ± standard deviation (SD) for normally distributed data or median (interquartile range (IQR)) for skewed distributions, and were compared using one-way analysis of variance or Kruskal-Wallis tests, respectively. Categorical variables are expressed as frequencies (percentages) and compared using χ^2^ tests.

Associations between GlycA and CMR-derived IDPs or ECG parameters were examined using multivariable linear regression models adjusted for potential confounders. Protein-GlycA associations were analyzed using the same approach. Mediation analyses investigated each protein as a potential mediator of GlycA-IDP associations. Specifically, we used the mediation package in R, considering each IDP as outcome variable, GlycA as predictor and proteins as mediators, and adjusted for potential confounders. Results are reported as average causal mediation effect (ACME), representing the average change in the outcome if the treatment were maintained constant but the mediator were altered, and mediated proportions, representing the ratio of ACME to the total effect, with 95% confidence interval (CI).

For survival analyses, participants were stratified by GlycA quintiles (Q1: lowest; Q5: highest). Cox proportional hazards models were used to test GlycA as an independent predictor of primary and secondary endpoints beyond CMR features or lipid profiles, with adjustment for potential confounders. Sensitivity analyses employed Fine-Gray subdistribution hazards regression with non-cardiovascular death as a competing risk. Non-linear associations were explored using restricted cubic splines. Results are reported as hazard ratio (HR) with 95% CI. Mediation analyses examined proteins as potential mediators of GlycA-incident MACE associations using parametric survival models with Weibull distribution, adjusted for confounders and lipid profiles.

Given that hsCRP represents the most widely studied inflammatory biomarker parallel analyses were conducted substituting hsCRP for GlycA.

Statistical significance was defined as *P* < 0.05 (two-sided). Multiple comparison correction was performed using the Benjamini-Hochberg false discovery rate method.^26^

## Results

### Study cohort

A total of 502,128 participants aged 40–69 years were enrolled across the United Kingdom between 2006 and 2010. Of these, 189 individuals were excluded due to withdrawn consent, leaving 501,939 participants eligible for analysis.

Of the 501,936 participants with GlycA measurements, 488,079 individuals remained after metabolomic data preprocessing. The effect of chronic inflammation on cardiac remodeling was evaluated in participants with both CMR (n = 81,313) and ECG (n = 81,964) data, yielding 70,809 individuals with complete GlycA, CMR, and ECG measurements. Protein mediation analyses examining the interaction between GlycA and IDPs, as well as between GlycA and MACE, were conducted in 8,184 and 48,427 participants with available proteomic data, respectively.

For survival analyses, participants with MACE occurring before CMR imaging (n = 45,733) were excluded when testing inflammation effects on outcomes adjusted for IDPs, whereas those with MACE before enrollment (n = 19,131) were excluded when testing inflammation effects adjusted for lipid profiles. This resulted in 72,541 and 454,821 individuals for CMR-adjusted and lipid-adjusted analyses, respectively.

For XWAS analyses, environmental exposure data were available for 492,110 individuals. After excluding 13,169 participants with incomplete data, 478,941 individuals had complete GlycA and environmental exposure measurements. Sample sizes for body composition analyses were detailed above.

Gene-environment interactions were investigated using 10 multi-ancestry PRSs for cardiovascular disease, which were available for 485,717 individuals.

### Inflammation-driven cardiac remodeling

Baseline characteristics of participants with available metabolomic data are shown in Table 1. The majority of participants were women (54%) and of self-reported white ethnicity (95%). At recruitment, mean age was 57±8 years, with 27% having hypertension, 10% being current smokers, 5.3% having diabetes, and 7% having previous MACE. Mean GlycA concentration was 0.82±0.12 mmol/L for both sexes, and significantly differed among ethnicities, with afro-caraibbean individuals showing the lowest value (all *P* < 0.001). Supplementary Table 6 shows demographic characteristics and IDPs stratified by GlycA quintiles. Body mass index, age, and blood pressure increased progressively across quintiles (*P* < 0.001 for all) (Figure 2).

**Figure 2.**
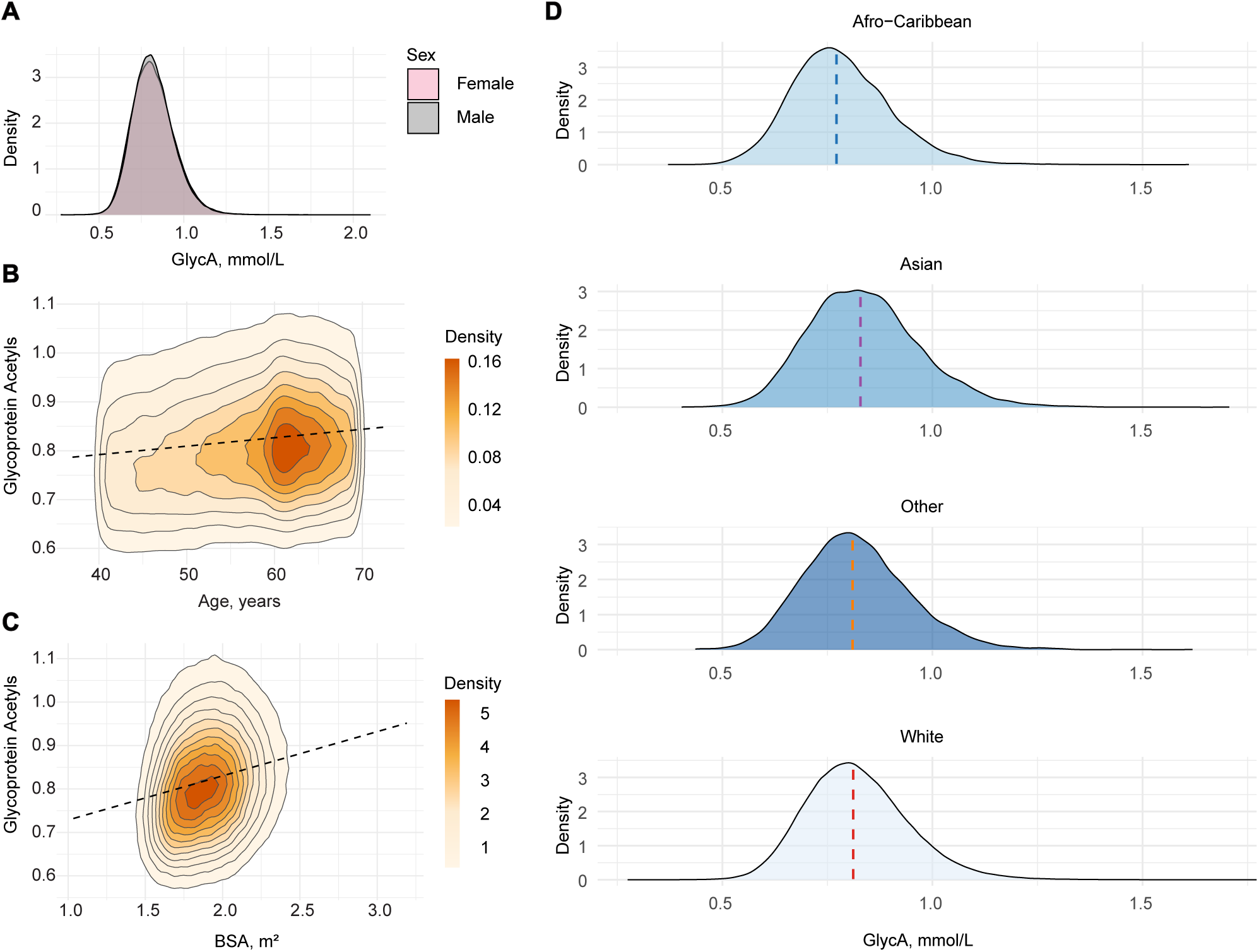
Distribution of glycoprotein acetyls values. **a)** Ridge plots summarising the distribution densities of GlycA in men and women subgroups. Unadjusted values shown. **b,c)** Scatter plots of GlycA with age **(b)** and BSA **(c)**, with linear model fit and marginal density plots. **d)** Ridge plots summarising the distribution densities of GlycA across ethnicities. Unadjusted values shown. **Abbreviations:** BSA, body surface area; GlycA, glycoprotein acetyls.

**Table 1.**
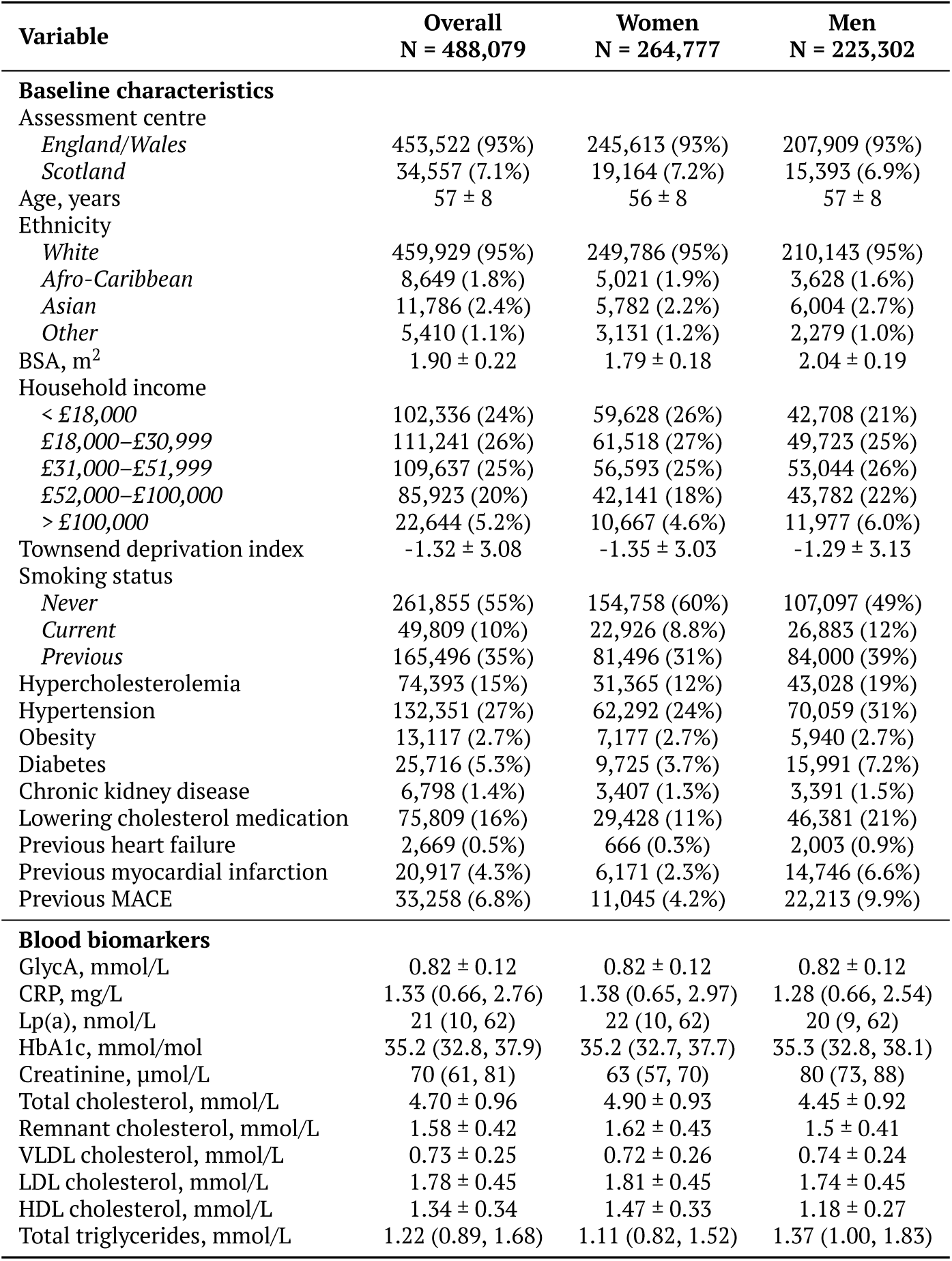
Baseline characteristics. Demographic, risk factors and blood biomarker among participants with available metabolomic data.

After adjusting for confounders, higher GlycA was associated with reduced volumes across all cardiac chambers, e.g. left ventricle (LV) EDVi (β = −2.09, *P* < 10^−200^) (Figure 3a), as well as increased global LV wall thickness, reduced indexed LV mass (β = −0.30, *P* < 10^−20^) and reduced diastolic function (longitudinal PDSR: β = −0.04, *P* < 10^−90^). Furthermore, higher GlycA was associated with reduced stroke volume and increased heart rate (β = 1.38, *P* < 10^−200^), but a negligible effect on cardiac index (β < 0.01) (Supplementary File 1).

**Figure 3.**
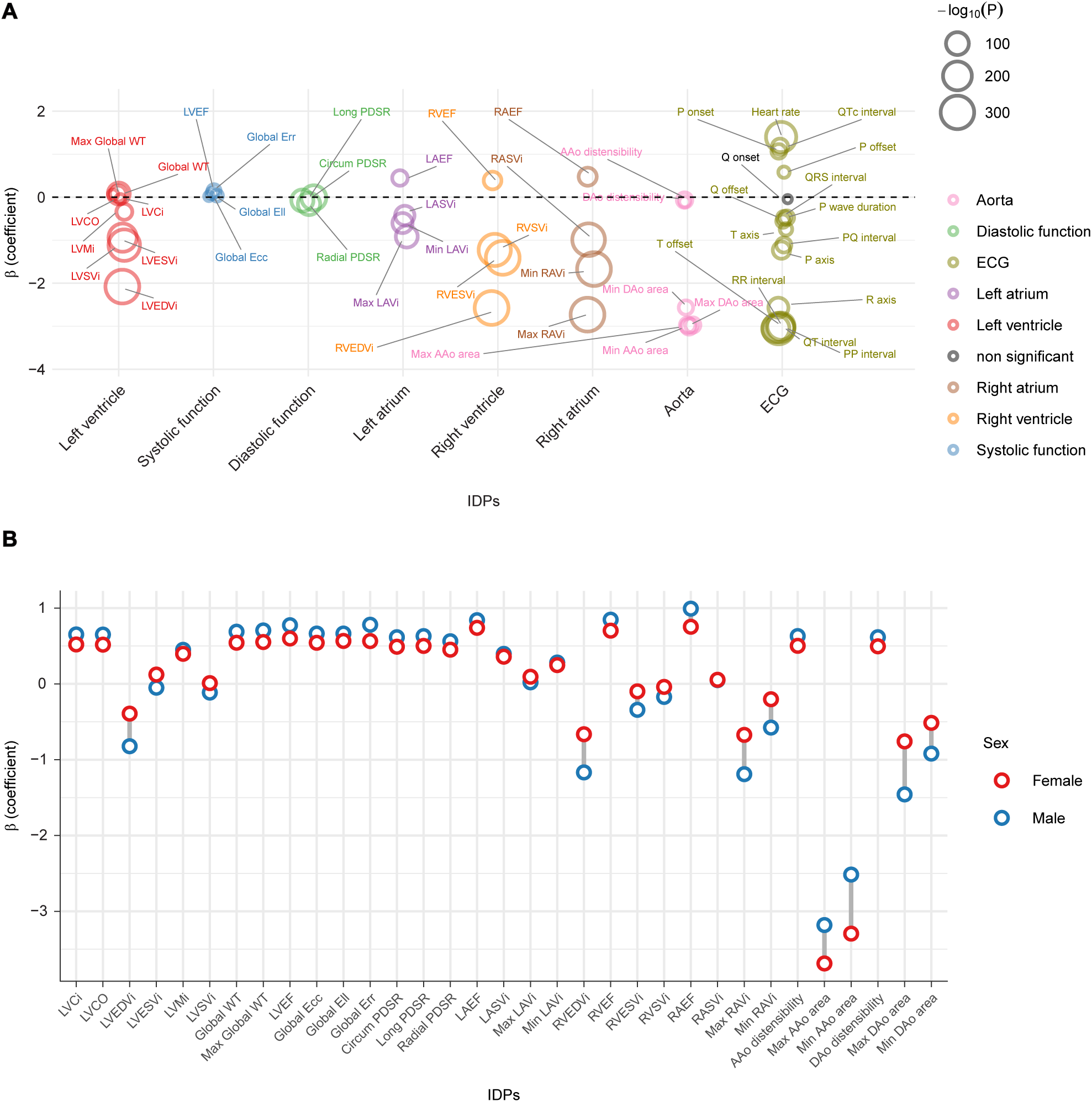
Effect of chronic inflammation on cardiac geometry. **a)** Multivariable linear regression coefficients for the effect of GlycA on IDPs and ECG variables in the pooled cohort. Each IDP or ECG variable is considered as predictor, and regressed against GlycA after adjusting for BSA, age at the date of CMR, sex, age:sex interaction, age^2^, and diabetes status. β coefficients are scaled and a minimum cap of −3 is set for clarity; horizontal and vertical jitters are added to reduce points overlap. **b)** Multivariable linear regression coefficients for the effect of GlycA on IDPs stratified by sex. β coefficients are scaled. **Abbreviations:** AAo, ascending aorta; BSA, body surface area; CMR, cardiac magnetic resonance; DAo, descending aorta; DBP, diastolic blood pressure; ECG, electrocardiogram; EDVi, end-diastolic volume indexed; Ell, longitudinal strain; Ecc, circumferential strain; Err, radial strain; ESVi, end-systolic volume indexed; GlycA, glycoprotein acetyl; IDP, imaging-derived phenotype; LA, left atrium; LAEF, left atrial ejection fraction; LASVi, left atrial stroke volume indexed; LAVmax, maximum left atrial volume indexed; LAVmin, minimum left atrial volume indexed; LV, left ventricle; LVCO, left ventricular cardiac output; LVCi, left ventricular cardiac index; LVEDVi, left ventricular end-diastolic volume indexed; LVEF, left ventricular ejection fraction; LVESVi, left ventricular end-systolic volume indexed; LVMi, left ventricular mass indexed; LVSVi, left ventricular stroke volume indexed; MAP, mean arterial pressure; PDSR, peak diastolic strain rate; P, P wave; Q, Q wave; QRS, QRS interval; R, R wave; RA, right atrium; RAEF, right atrial ejection fraction; RASVi, right atrial stroke volume indexed; RAVmax, maximum right atrial volume indexed; RAVmin, minimum right atrial volume indexed; RV, right ventricle; RVEDVi, right ventricular end-diastolic volume indexed; RVEF, right ventricular ejection fraction; RVESVi, right ventricular end-systolic volume indexed; RVSVi, right ventricular stroke volume indexed; SBP, systolic blood pressure; SVi, stroke volume indexed; T, T wave; WT, myocardial wall thickness.

Findings were consistent across sex-stratified analyses (Figure 3b; Supplementary Files 2-3), diabetes status (r = 0.985, *P* = 3.1 × 10^−37^; Supplementary Figure 1a; Supplementary Files 4-5), obesity status (r = 0.993, *P* = 8.7 × 10^−46^; Supplementary Figure 1b; Supplementary Files 6-7), and age groups (Supplementary Files 8-10). Associations persisted after adjusting for hsCRP (Supplementary File 11).

We next investigated the role of 80 inflammatory proteins in cardiac remodeling. First, we identified proteins significantly associated with inflammatory biomarkers (both GlycA and hsCRP), as reported in Supplementary Figure 2 and Supplementary Files 12-13. Second, we assessed whether these proteins mediate the association between GlycA and cardiac remodeling. As shown in Figure 4a, numerous proteins significantly mediated effects on LV and right ventricle (RV) EDVi, ESVi, and atrial volumes, predominantly involving tumor necrosis factor (TNF), transforming growth factor, Interleukin (IL)-1, and IL-6 signaling pathways. Figure 4b-e illustrates the proportion of mediated effect for four IDPs reflecting LV geometry.

**Figure 4.**
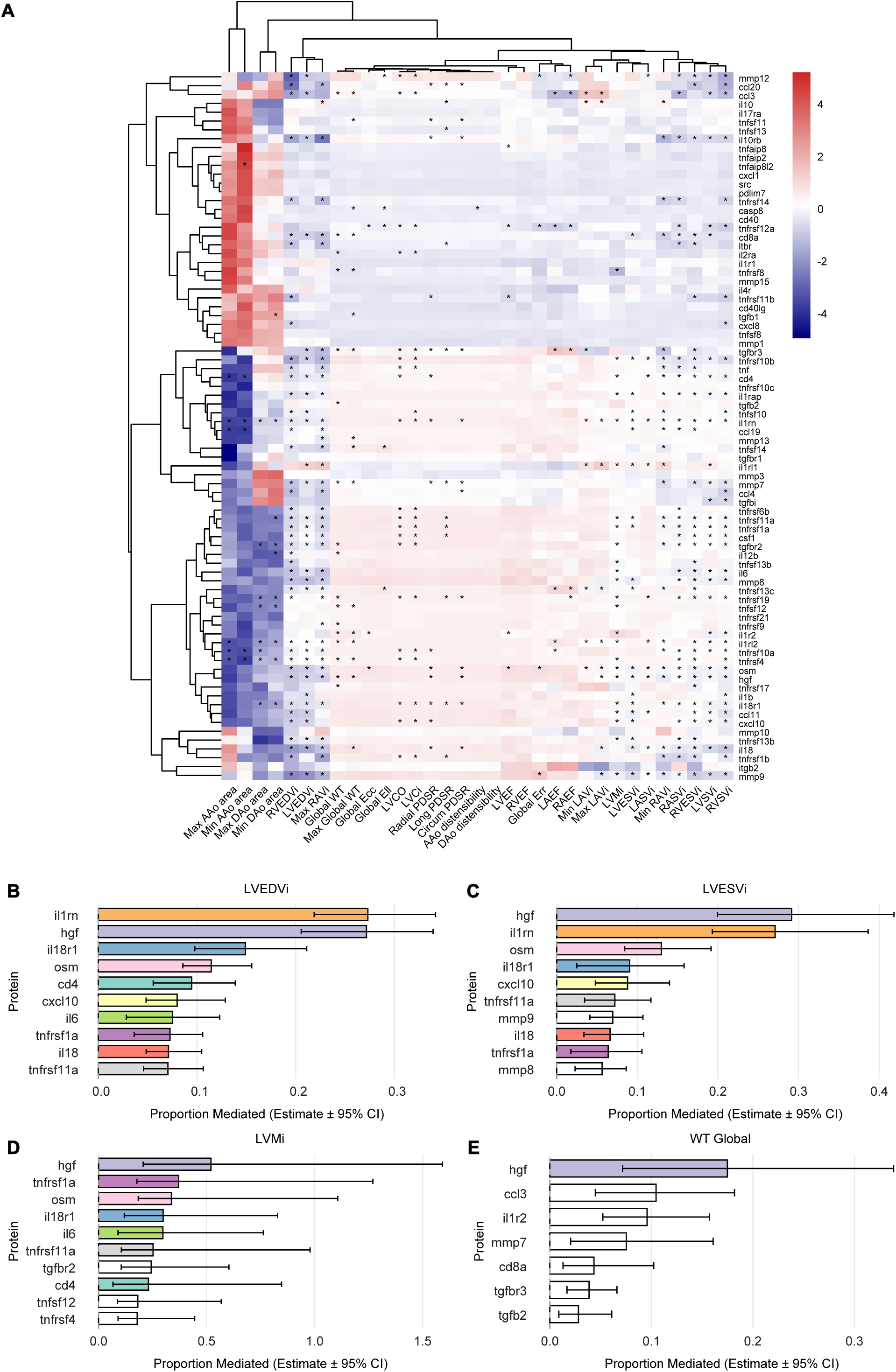
Proteins mediating the effect of inflammation on cardiac geometry. **a)** Heatmap showing the scaled coefficients for the average causal mediation effect of each protein on each IDP. Each model included log-transformed and scaled GlycA as the exposure, one protein as the mediator, and one IDP as the outcome. Models were adjusted for BSA, sex, age, age^2^, the age–sex interaction, and prevalent diabetes. (*) denotes an FDR-adjusted p-value < 0.05. The bottom panel reports the proportion of the mediated effect for the top 10 proteins for LVEDVi (**b**), LVESVi (**c**), LVMi (**d**), and global WT (**e**). **Abbreviations:** AAo, ascending aorta; BSA, body surface area; DAo, descending aorta; EDVi, end-diastolic volume indexed; Ell, longitudinal strain; Ecc, circumferential strain; Err, radial strain; ESVi, end-systolic volume indexed; GlycA, glycoprotein acetyl; IDP, imaging-derived phenotype; LA, left atrium; LAEF, left atrial ejection fraction; LASVi, left atrial stroke volume indexed; LAVmax, maximum left atrial volume indexed; LAVmin, minimum left atrial volume indexed; LV, left ventricle; LVCO, left ventricular cardiac output; LVCi, left ventricular cardiac index; LVEDVi, left ventricular end-diastolic volume indexed; LVEF, left ventricular ejection fraction; LVESVi, left ventricular end-systolic volume indexed; LVMi, left ventricular mass indexed; LVSVi, left ventricular stroke volume indexed; MAP, mean arterial pressure; PDSR, peak diastolic strain rate; RA, right atrium; RAEF, right atrial ejection fraction; RASVi, right atrial stroke volume indexed; RAVmax, maximum right atrial volume indexed; RAVmin, minimum right atrial volume indexed; RV, right ventricle; RVEDVi, right ventricular end-diastolic volume indexed; RVEF, right ventricular ejection fraction; RVESVi, right ventricular end-systolic volume indexed; RVSVi, right ventricular stroke volume indexed; SVi, stroke volume indexed; WT, myocardial wall thickness.

Interleukin-1 receptor antagonist protein (IL-1RA) mediated the largest proportion of the GlycA effect on LV EDVi (ACME −0.53 [95% CI: −0.64 to −0.41, *P* < 10^−16^], proportion mediated 0.27 [95% CI: 0.22–0.34, *P* < 10^−16^]) and demonstrated the strongest ACME on indexed LV mass (−0.18 [95% CI: −0.24 to −0.11, *P* < 10^−16^]). Hepatocyte growth factor (HGF) mediated the largest proportion of the GlycA effect on LV ESVi (ACME −0.26 [95% CI: −0.33 to −0.18, *P* < 10^−16^], proportion mediated 0.29 [95% CI: 0.20–0.42, *P* < 10^−16^]) and LV global wall thickness (ACME 0.01 [95% CI: 0.01 to 0.02, *P* < 10^−16^], proportion mediated 0.17 [95% CI: 0.06–0.34, *P* < 10^−16^]).

Extended data are provided in Supplementary File 14. Mediation analyses for hsCRP effects on IDPs yielded comparable results (Supplementary Figure 3 and Supplementary File 15).

### Outcome analysis

After a median follow-up of 15.3 (14.3-16.0) years (17,777 person-years), 74,336 individuals experienced the primary endpoint. The majority of MACE events were myocardial infarction (7.7%), followed by stroke (6.6%) and HF (4.3%). For the secondary endpoint, 55,076 (11%) all-cause deaths were observed, including 10,377 cardiovascular deaths (Supplementary Table 7).

Covariates (BSA, sex, age, age^2^, age:sex interaction, and statin use), lipid, and competing risk-adjusted cumulative incidence curves for the probability of incident MACE and all-cause death rose with each quintile of GlycA (Figure 5). The covariate-adjusted risk for first MACE across each quintile of GlycA is shown in Supplementary Table 8. The HR for the primary endpoint comparing quintile 5 with quintile 1 was 1.64 (1.60-1.69) for the covariate-only adjusted model, 1.46 (1.41-1.52) for the covariate plus lipid profile model, and 1.42 (1.38-1.49) for the competing risk model. After adjusting the competing risk model for hsCRP, HR declined to 1.14 (1.09-1.18). Finally, GlycA significantly predicted the outcome independently of IDPs, with a HR of 1.13 (1.08–1.19) per SD increase. Restricted cubic spline analysis showed a nonlinear association between GlycA and MACE, with risk remaining stable at lower levels and increasing markedly above the median (*P* < 0.001 for nonlinearity) (Supplementary Figure 4).

**Figure 5.**
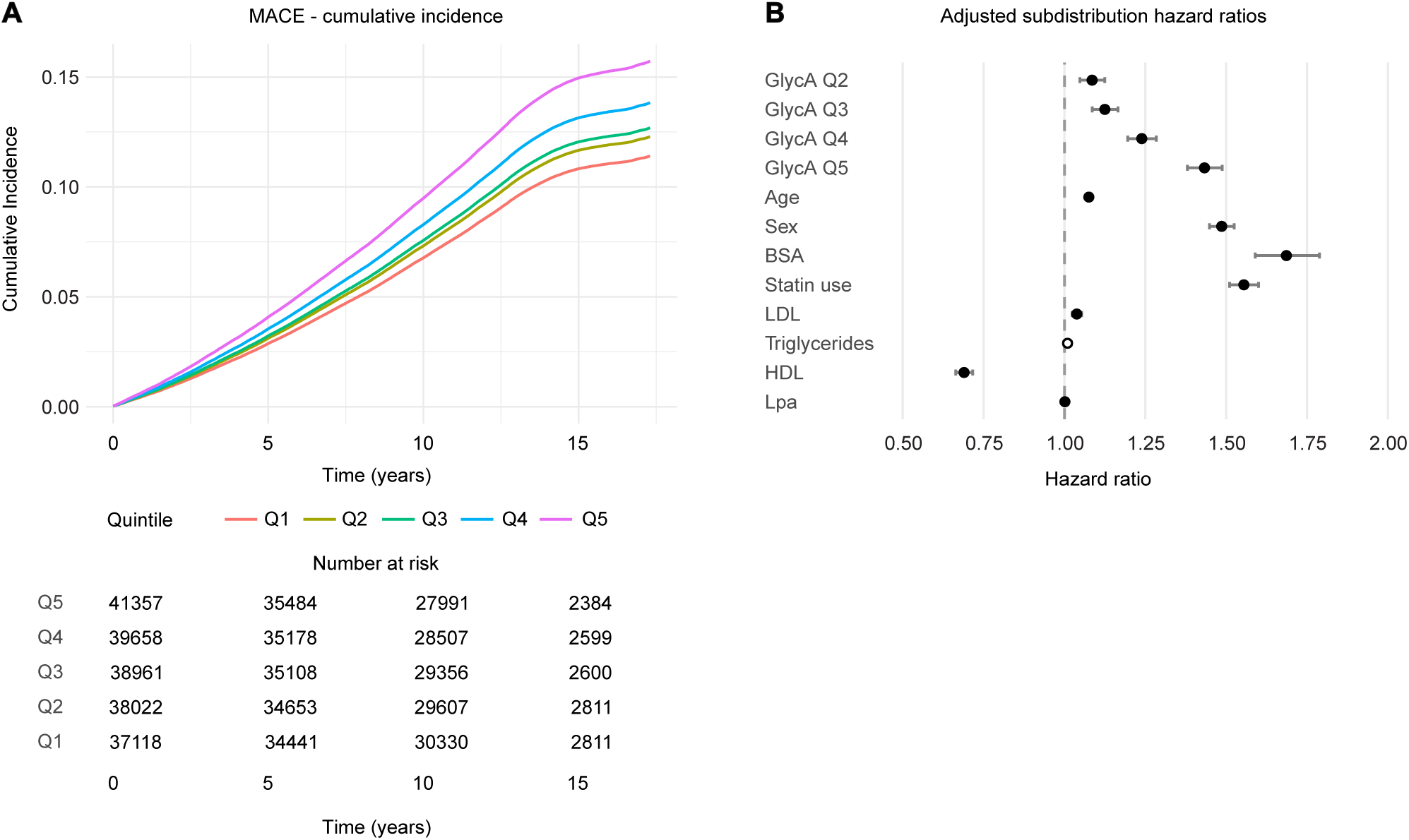
Cumulative incidence of first major cardiovascular events. **a)** Covariate- and competing risk–adjusted cumulative incidence of first major cardiovascular events among initially healthy participants, stratified by quintiles of baseline GlycA levels. **b)** Forest plot showing adjusted subdistribution hazard ratios from covariate- and competing risk–adjusted models across quintiles of baseline GlycA levels. Quintile 1 included the lowest biomarker levels and quintile 5 the highest levels. **Abbreviations:**GlycA, glycoprotein acetyls; MACE, major adverse cardiovascular event.

Similar results were found using log-transformed and scaled hsCRP instead of GlycA (Supplementary Table 9), and when considering all-cause death as the outcome variable (Supplementary Table 10).

To further investigate inflammatory proteins mediating the effect of GlycA on incident MACE, we conducted mediation analysis using the proteomic data. Sixty-three proteins significantly mediated the association between GlycA and MACE (Figure 6a). Among the 20 proteins with the largest mediated effects, those most represented belonged to the TNF and IL-6 signaling pathways. Figure 6b shows the proportion of mediated effect for the 20 proteins with the highest absolute beta coefficients, where negative values indicate mediation of an association with shorter time-to-event.

**Figure 6.**
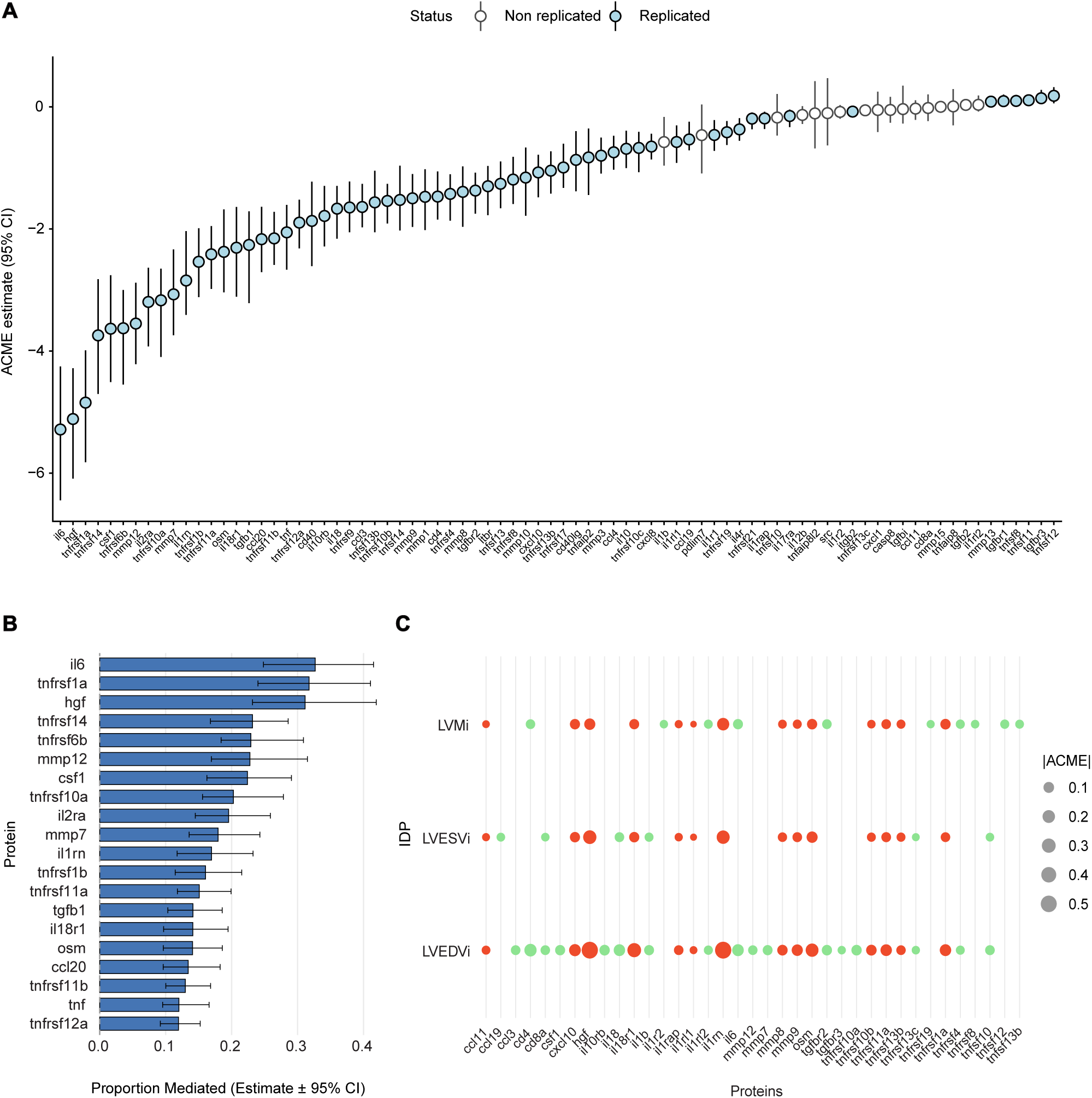
Proteins mediating the effect of inflammation on incident MACE. **a)** Average causal mediation effect and corresponding 95% CI for each protein. Each model assessed one protein as a potential mediator of the association between GlycA and incident MACE. A parametric survival model assuming a Weibull distribution was used, and models were adjusted for BSA, sex, age, age^2^, the age–sex interaction, statin use, LDL cholesterol, triglycerides, HDL cholesterol, and Lp(a). **b)** Top 20 proteins with the highest proportion of mediated effect. **c)** Proteins significantly mediating the effect of inflammation on survival and on LVEDVi, LVESVi, and LVMi. Red dots indicate proteins that replicated across all three IDPs and the survival analysis. Dots size is proportional to the absolute ACME value. **Abbreviations:** ACME: average causal mediation effect; BSA, body surface area; HDL, high-density lipoprotein; LDL, low-density lipoprotein; Lp(a), lipoprotein a; IDP, imaging-derived phenotype; LVEDVi, left ventricle end-diastolic volume; LVESVi, left ventricle end-systolic volume; LVMi, left ventricle mass index; MACE, major adverse cardiovascular event.

IL-6 exhibited the largest mediated effect of GlycA on incident MACE (ACME −5.28 [95% CI: −6.44 to −4.25, *P* < 10^−16^]; proportion mediated 0.33 [95% CI: 0.25-0.41, *P* < 10^−16^]), followed by HGF (ACME −5.11 [95% CI: −6.09 to −4.28, *P* < 10^−16^]; proportion mediated 0.31 [95% CI: 0.23-0.42, *P* < 10^−16^]). Notably, IL-1RA and HGF—both of which showed strong mediation effects on LV remodeling—also strongly mediated the association with the primary endpoint (Supplementary Files 16-17). Figure 6c shows proteins that significantly mediate the association between GlycA, incident MACE, LV EDVi, LV ESVi, and indexed LV mass.

### Exposures predicting inflammation

XWAS was conducted on 478,941 individuals with environmental factors and GlycA data available, and on 70,528 participants with available body fat composition measurements, testing 177 exposures in total (Supplementary File 18).

In all participants 166 of 177 variables were significantly associated with GlycA levels (Figure 7a, Supplementary File 19). Body fat composition, smoking status, psychological conditions, and low socioeconomic status showed the strongest associations with GlycA after adjusting for confounders. Total trunk fat mass (β = 0.35, *P* < 10^−50^), current smoking status (β = 0.39, *P* < 10^−50^), and renting from local authorities (β = 0.36, *P* < 10^−50^)—a marker of low socioeconomic status—showed the strongest associations with inflammatory levels. Depressive symptoms, visceral fat and android-distribution fat were also strongly positively associated with chronic inflammation.

**Figure 7.**
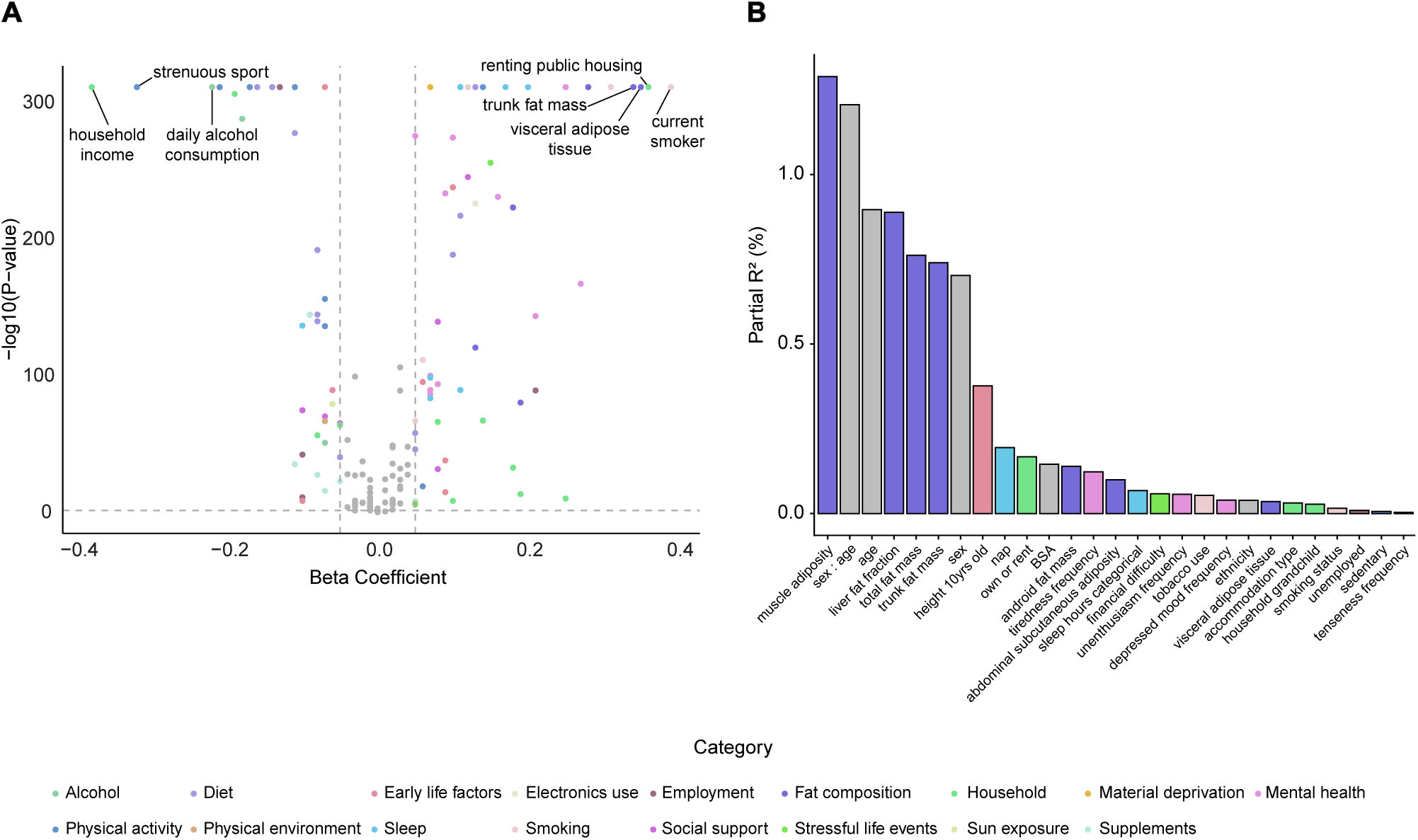
Exposome-wide association study. **a)** Volcano plot showing the − log_10_ FDR-adjusted *P* value of the β coefficient for the association between each exposure (predictor) and GlycA (outcome). Quantitative exposure variables were log-transformed and standardized. Models were adjusted for BSA, sex, age, age^2^, age:sex interaction, and ethnicity. Grey dots indicate exposures with an absolute β < 0.05. Exposures with the largest absolute β values are labeled. **b)** Bar plot showing the partial *R*^2^ of each exposure from a multivariable linear model with GlycA as the outcome, including the 20 exposures with the largest absolute β values and adjusted for BSA, sex, age, age^2^, age:sex interaction, and ethnicity. **Abbreviations:** BSA, body surface area; FDR, false discovery rate; GlycA, glycoprotein acetyls.

In contrast, variables reflecting higher socioeconomic status, physical activity, and healthier diet were negatively associated with GlycA levels. Household income and engaging in strenuous exercise four times per week showed the strongest negative associations (*β* = −0.38 and −0.32, respectively, *P* < 10^−50^ for both). Oily fish and total fruit consumption were also significantly associated with lower inflammation levels. When the 25 exposures most strongly associated with GlycA were included in a multivariable linear model together with confounders, the full model explained 18.6% of the variance in GlycA. Individually, however, most exposures contributed only a small proportion of the explained variance (Figure 7b).

No notable differences were observed in XWAS regression coefficients when analyses were performed separately by sex (Spearman correlation coefficient r = 0.937, *P* < 10^−50^; Supplementary Figure 5a, Supplementary Files 20-21). Similarly, a strong correlation was observed between regression coefficients when hsCRP was used instead of GlycA as the predictor (Spearman correlation coefficient r = 0.926, *P* < 10^−50^; Supplementary Figure 5b, Supplementary File 22).

### Gene-environment interactions on inflammation

We investigated interactions between multi-ancestry PRSs and environmental factors in determining inflammatory levels in 485,717 individuals. We identified 115 exposure–PRS interactions that significantly modified the relationship between environmental factors (as predictors) and GlycA (as the outcome) (Supplementary File 23). After further adjusting for the Townsend deprivation index — a marker of low socioeconomic status — 101 interactions remained significant (Figure 8a, Supplementary File 24). The type 2 diabetes PRS demonstrated the greatest number of significant interactions (35), while stroke, hypertension, and CVD PRSs each showed 13 significant interactions. Body fat composition, socioeconomic status, pollution, and mental health factors exhibited significant interactions with PRSs in determining GlycA levels. As an example, the association of depressed mood with GlycA was amplified with increasing genetic risk, ranging from the lowest type 2 diabetes PRS quintile (*β* = 0.19) to the highest quintile (*β* = 0.25).

**Figure 8.**
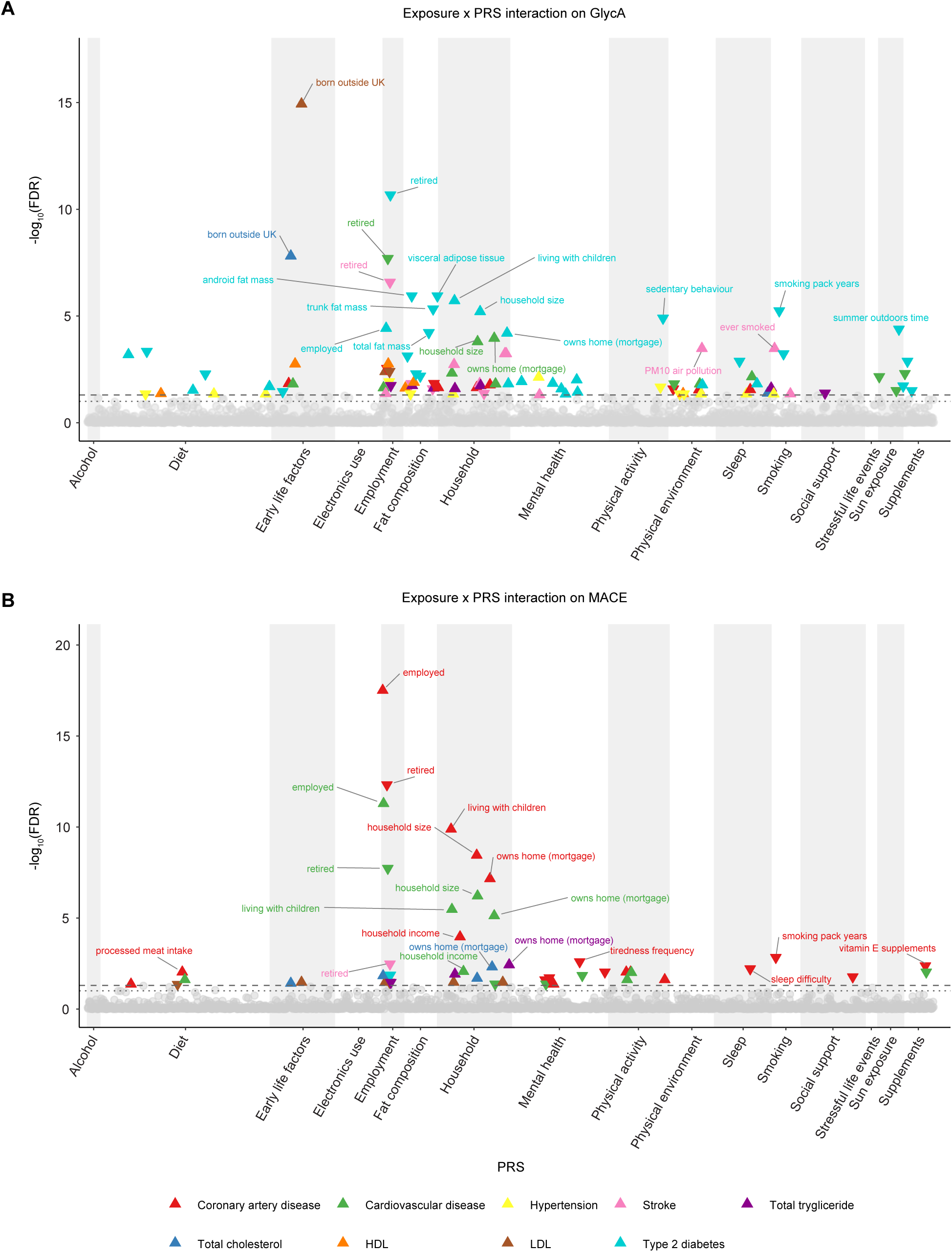
Exposure-gene interactions. **a)** Manhattan plot showing the − log_10_ FDR-corrected *P* values of the interaction terms between exposures and PRS in predicting GlycA levels. Models were fitted with log-transformed GlycA as the dependent variable and exposures as independent variables. For each exposure, all PRS were tested in separate models by including a multiplicative interaction term, and models were adjusted for BSA, sex, age, age^2^, age–sex interaction, ethnicity, and Townsend deprivation index. Triangles indicate interactions with an FDR-adjusted *P* < 0.05. The top 20 interactions are labeled. Non-significant interactions are shown as gray dots. Upward-pointing triangles denote positive interaction effects (β > 0), whereas downward-pointing triangles denote negative interaction effects (β < 0). **b)** Manhattan plot showing the − log_10_ FDR-corrected *p*-values of the interaction terms between exposures and PRS in predicting incident MACE. For each exposure, all PRS were tested in separate models by including a multiplicative interaction term, and models were adjusted for BSA, sex, age, age^2^, age–sex interaction, ethnicity, and Townsend deprivation index. Triangles indicate interactions with an FDR-adjusted *P* < 0.05. The top 20 interactions are labeled. Non-significant interactions are shown as gray dots. Upward-pointing triangles denote interaction effects with β > 1, whereas downward-pointing triangles denote interaction effects with β < 1. **Abbreviations:** FDR, false discovery rate; GlycA, glycoprotein acetyl; HDL, high-density lipoprotein; LDL, low density lipoprotein; MACE, major adverse cardiac event; PRS, polygenic risk score; UK; United Kingdom.

We observed 51 and 49 exposure–PRS interactions that significantly affected the association between GlycA levels and MACE before and after adjustment for the Townsend deprivation index, respectively (Figure 8b, Supplementary Files 25–26). Coronary artery disease and CVD PRSs demonstrated significant interactions with 20 and 13 exposures, respectively. Socioeconomic status, mental health, and physical activity emerged as the strongest exposure categories. Specifically, the association of employment with MACE was augmented with increasing genetic risk, ranging from the lowest CVD PRS quintile (HR = 0.79) to the highest quintile (HR = 0.85).

Notably, 17 exposure–PRS interactions were replicated across both analyses (GlycA and MACE), predominantly involving socioeconomic variables. For instance, employment status significantly interacted with the CVD PRS, amplifying its effect on both GlycA (*β* = 0.01, *P* = 0.02) and MACE (*β* = 1.07, *P* < 10^−10^).

## Discussion

Our findings demonstrate that chronic systemic inflammation associates with restrictive cardiac remodeling characterized by reduced ventricular chamber volumes, diastolic impairment, and a compensatory chronotropic response. Circulating proteins, including IL-1, emerged as key mediators of structural cardiac adaptation and adverse clinical outcomes. Elevated GlycA also independently predicted incident cardiovascular events and all-cause mortality beyond traditional risk factors and lipid levels. Additionally, we identified significant gene-environment interactions in which genetic susceptibility modified the inflammatory response to environmental exposures, suggesting that individual risk is determined by the convergence of inherited and acquired factors.

### GlycA as a cardiovascular risk biomarker

We considered GlycA, an NMR spectroscopy–derived signal quantifying the circulating levels of multiple immunoregulatory proteins, as a marker of chronic inflammation. It has emerged as a stronger inflammatory biomarker compared to hsCRP,^28^ as it reflects the activation of multiple biological pathways,^29^ exhibits lower long-term intra-individual variability,^30^ and is less sensitive to ethnicity, sex and renal function.^31–33^ GlycA has also been associated with incident CVD in smaller observational studies, making it an attractive systemic inflammatory biomarker for cardiovascular risk assessment.^12,34–37^

Our study represents the largest analysis to date, with long-term follow-up, assessing the relevance of GlycA as a cardiovascular risk factor. Specifically, we investigated the association between GlycA and adverse cardiac events in a large cohort of approximately 480,000 individuals, performing sensitivity analyses adjusting for potential confounders, established biomarkers, and competing risks. Our results support GlycA as a clinically relevant predictor of MACE in a community population, aligning with findings observed in smaller primary prevention cohorts. GlycA correlated with hsCRP levels but provided incremental prognostic information consistent with “sensing” complementary inflammatory risk pathways that mediate outcomes and being a more robust predictor of cytokines.^38,39^

### Inflammation and cardiac remodeling

Systemic inflammation has been recognized as a key pathobiologic driver of the development, progression, and complications of many CVDs, including atherosclerosis, myocarditis, and HF.^40^ Activation of inflammatory pathways has been implicated in the hemodynamic progression of heart failure with reduced ejection fraction (HFrEF) and heart failure with preserved ejection fraction (HFpEF).^16,41–43^ While acute inflammation, assessed using hsCRP, has been linked to incident HF,^34^ the pattern of remodeling that precedes overt disease and how this is mediated over time in the population by circulating factors is unknown.

Here, we investigated this relationship in a large community-living cohort with multi-omic profiling and CMR. We found that chronic inflammation was associated with restrictive remodeling characterized by reduced chamber volumes and LV mass, increased wall thickness, and mild diastolic impairment. The absence of atrial dilation suggests that filling pressures may not be chronically elevated. A modest increase in heart rate likely reflects physiological compensation for reduced stroke volumes. This phenotype is similar to that seen in uncomplicated diabetes,^44^ but in our study prevalent diabetes did not modify the inflammation-remodeling association. Potential mechanisms of inflammasome-mediated restrictive remodeling include increased extracellular matrix deposition with up-regulation of collagen expression as well as impaired calcium homeostasis and depressed myocyte contractility.^45^

### Proteomic mediators as therapeutic targets

We investigated 80 circulating proteins identifying 14 biomarker candidates associated with restrictive remodeling and incident MACE which were directly associated with GlycA and hsCRP levels. The most represented pathways involved the IL-1 and TNF superfamilies. IL-1RA, an endogenous inhibitor of IL-1α/*β* activity,^46,47^ has been positively associated with cardiovascular risk,^47^ although IL-1 blockade has failed to improve outcomes in obese HFpEF patients.^48^ However, IL-18, another IL-1 axis mediator and critical factor in so-called auto-inflammatory IL-18opathies,^49^ shows promise: receptor knockout mice exhibit attenuated LV dysfunction,^50,51^ genetic variation appears to influence cardiovascular outcomes,^52,53^ and pathway inhibition is currently being tested in clinical studies.^54,55^

Within the TNF superfamily, TNFrsf1A predicts HFpEF development and mortality,^56^ although anti-cytokine therapy has not been successful in HFrEF trials.^57^ TNFrsf11A (RANK/RANKL axis) and TNFrsf13B (B-cell immunity) are associated with CVD^58,59^ and are therapeutic targets in other inflammatory conditions.^60,61^ IL-6’s association with MACE and LVEDVi is particularly relevant given ongoing trials of the monoclonal antibody ziltivekimab targeting residual inflammatory risk.^62^ Additional mediators—HGF, matrix metalloproteinases 8 and 9, oncostatin M, and C-X-C motif chemokine ligand 10—represent further potential therapeutic targets emerging from our data.

### Environmental exposures and genetic susceptibility

Our study provides the first large-scale systematic assessment of the relative contributions of environmental exposures and genetic susceptibility on chronic inflammation, as well as their interaction, in a population-based setting. Previous epidemiological studies have demonstrated robust associations between individual environmental and behavioral exposures and aging or mortality using exposome-wide approaches;^7^ however, comparable comprehensive analyses focusing on inflammatory phenotypes have been lacking. In this context, our XWAS extends evidence from smaller observational studies, identifying smoking behavior, body fat composition, mental health, and socioeconomic status as the environmental factors most strongly associated with chronic inflammation.^63–66^

Beyond the main exposure effects, we systematically evaluated interactions between environmental exposures and polygenic risk for a range of CVDs, demonstrating that genetic susceptibility significantly modifies the relationship between environmental risk burden and inflammation. These findings indicate that individual inflammatory responses to environmental exposures are shaped by inherited risk and that polygenic scores for multiple CVDs may act through modulation of shared inflammatory pathways. Future work might examine whether combining polygenic risk assessment with biomarkers of chronic inflammation could predict susceptibility or resilience to inflammatory stressors and identify those most at risk of poor outcomes associated with modifiable cardiovascular risk factors and other environmental exposures.

### Limitations

The UKB population is primarily of European descent highlighting the need for enriching future analyses with more diverse populations. Participants in the imaging sub-study were invited at random,^67^ but while the population is subject to healthy volunteer selection bias it still enables valid scientific inferences of associations between exposures, genetic variation and health conditions that are generalizable.^68^ GlycA is assessed with NMR which is not currently available in routine healthcare settings and its predictive value compared to hsCRP needs to be validated in broader populations.

Although a relatively large number of proteins were examined, they represent only a subset of the circulating proteome and do not necessarily capture intracellular or tissue-specific protein expression. Environmental exposures were measured with varying precision and do not capture the full complexity of the exposome, introducing potential measurement error and residual confounding. As example, self-reported alcohol intake in the UKB has been shown to be subject to measurement error.^69^ Lastly, mediation analyses conducted in a cross-sectional setting should be interpreted with caution, as the temporal ordering of exposure, mediator, and outcome cannot be established and associations may be bidirectional.

## Conclusions

These findings advance understanding of the role of chronic inflammation in the etiology of heart disease, highlighting inflammatory pathways as key contributors to adverse cardiac remodeling and cardiovascular risk and revealing exposure-gene interactions as critical modulators of individual inflammatory responses.

## Funding

The study was supported by the Medical Research Council (MC_UP_1605/13); British Heart Foundation (RG/F/24/110138, RE/24/130023, CH/F/24/90015); and the National Institute for Health Research (NIHR) Imperial College Biomedical Research Centre. D.P.O’R. is also supported by the British Heart Foundation’s Big Beat Challenge award to CureHeart (BBC/F/21/220106).

## Disclosures

D.P.O’R. has received fees from Bayer AG, and grant funding from Bayer AG and Calico Labs.

## Author contributions

Conceptualization: M.C., D.O.R.; Methodology: M.C., D.O.R., S.T., K.R.; Formal analysis: M.C., L.H., S.K., P.G., J.Z.; Writing - original draft: M.C.; Writing - review & editing: L.H., S.K., L.C., D.O.R.; Visualization: M.C., D.O.R.; Supervision: D.O.R; Project administration: D.O.R.; Funding acquisition: D.O.R.

## Data Availability

Data from UK Biobank are available for approved research.

## Notes

### Author Declarations

The study received ethical approval from the National Research Ethics Service (11/NW/0382), and all participants gave written informed consent.

