## Supplementary Materials for "Gene-exposure interactions regulate cytokine-mediated chronic inflammation and cardiac remodeling"

### Supplementary Material

### Supplementary Methods

#### Metabolomics

UKB metabolomics consisted of 249 metabolites measured from 501,936 participants across baseline and repeat. Nightingale NMR completed the biomarker processing on non-fasting plasma samples, the full methods are described elsewhere.[18] Metabolomic data was extracted using the Table Exporter tool using fields codes for categories 220, 221 and 222 on the UKB Showcase, obtaining 168 absolute and 81 derived metabolite measures. Pre-processing was performed to remove technical variation across both instances using the `ukbnmr` R package,[70], resulting in processed measures for 108 non-derived metabolites. Missing data for these 108 metabolites were then imputed for each instance using the `missRanger` R package, where participants with more than 50% and metabolites with more than 20% of data missing were removed prior to imputation. Next, the `ukbnmr` package was used to recalculate the derived measures from the 108 metabolites, resulting in a final imputed datasets of 325 metabolites for 488,079 participants. Further pre-processing steps included removal of outliers 6 SD deviations from the mean, log transformation and standardisation to mean 0 and SD 1.[71]

#### Exposure factors

The final list of the derived variables used for the XWAS analyses is provided in Supplementary File 19.

**Variable coding:** Variable recoding was performed before multiple imputation. All variable responses of "Prefer not to answer", "Do not know", and "Not applicable" were recoded to NA. Following multiple imputation, any variable values where the participant responded "Prefer not to answer" in the original dataset were recoded to NA. All nominal categorical variables were coded as unordered factors, with the reference level set as the most frequent response reported in the UK Biobank dataset. Ordered categorical variables were coded with the reference level set as the lowest response (e.g., "Never", "Rarely", or "None"). Dichotomous categorical variables were coded with the reference level set as "No." All variables with "mark all that apply" response categories were converted into multiple dummy variables, with each unique response option used to create a yes/no dichotomous variable.

For all numeric diet intake variables and number of hours spent driving, on the computer, or watching TV, participants who responded "less than one" were recoded as 0.5 hours. The "major dietary changes in the last 5 years" (field ID 1538) variable was recoded to NA if anyone reported "Yes, because of illness".

The home area population density variable (field ID 20118) was recoded with a classification of urban or rural home area population density. Participants who were classified in the original variable as "England/Wales – Urban – sparse," "England/Wales – Urban – less sparse," "Scotland – Large Urban Area," or "Scotland – Other Urban Area" were recoded to "Urban." All other responses were recoded to "Rural," with the exception of "Postcode not linkable" that was set as NA after imputation.

For smoking pack years (field ID 20161) and smoking pack years as a proportion of life span exposed to smoking (field ID 20162), missing values were recoded as 0 if the respondent was coded as "No" in response to the derived ever smoked variable available in the UKB (field ID 20160), but otherwise left as NA if respondents had ever smoked.

The "type of accommodation lived in" (field ID 670) was recoded to NA if anyone reported living in "sheltered accommodation".

**Recoding after imputation:** Several variables were recoded after multiple imputation. For bread type, cereal type, and coffee type variables, all participants with a response of 0 for the bread, cereal, and coffee intake questions were coded to "Never eat bread", "Never eat cereal," and "Never drink coffee," respectively. The "number of people living in your household" variable was recoded to NA if a respondent reported living in a care home or sheltered accommodation. Afterwards, dummy variables created from field ID 6141 (how are you related to people in your household) were recoded. Specifically, if the participant reported being the only person living in their household, then each dummy variable was set to "No." Additionally, if the participant reported living in a care home or sheltered accommodation, then each dummy variable was set to NA.

For processed meat (field ID 1349), poultry (field ID 1359), oily fish (field ID 1329), and non-oily fish (field ID 1339), the top three frequencies were combined to obtain four categories: never, <1.0 time per week, 1.0 time per week, and ≥2.0 times per week. For cheese intake (field ID 1408), the bottom two and the top two frequencies were combined to get four categories: <1.0 time per week, 1.0 time per week, 2.0–4.9 times per week, and ≥5.0 times per week. For daily tea intake (field ID 1488), participants were grouped as: <2.0 cups per day, 2.0–3.9 cups per day, 4.0–5.9 cups per day, and ≥6.0 cups per day. For daily coffee intake (field ID 1498), participants were grouped as: 0 cups/day, 0.5–1.9 cups per day, 2.0–2.9

cups per day, and ≥3.0 cups per day. Cereal (field ID 1458), bread (field ID 1448), and water (field ID 1528) intake were categorized into quartiles based on participants' responses. Cereal was coded as: <2 bowls/week, 2–4.9 bowls/week, 5–6.9 bowls/week, and ≥7 bowls/week. Bread was coded as: <8 slices/week, 8–13.9 slices/week, 14–19.9 slices/week, and ≥20 slices/week. Water was coded as: <1 glass/day, 1–1.9 glasses/day, 2–2.9 glasses/day, and ≥3 glasses/day.

For alcohol intake frequency (field ID 1558), all participants who responded as "Never" or "Previous" drinkers to the alcohol status variable (field ID 20117) were coded as NA. In addition, we coded participants who responded as drinking on "Special occasions only" as NA. The final variable was coded as a nominal variable with responses for "One to three times a month", "Once or twice a week", "Three or four times a week", and "Daily or almost daily", with "One to three times a month" set as the reference.

All derived variables were calculated after imputation, and the original variables used to create each of the derived variables were then excluded from the XWAS analysis.

**Sleep:** A categorical variable for hours of sleep was derived using the UKB hours of sleep continuous measure (field ID 1160). Category levels used for hours of sleep were: <7 hours, 7–9 hours, and >9 hours, with the reference set as 7–9 hours.[72]

**Bread and cereal fiber scores:** Scores were created using self-report data on intake of bread type, bread intake, breakfast cereal type, and breakfast cereal intake.[73] Bread and cereal intake, measured as a numeric response corresponding to portions consumed per week, were divided by 7 to get an estimate of daily intake, and then multiplied by the estimated fiber content for the specific type of bread and cereal that each participant reported to mainly eat. Continuous fiber scores were converted into quintiles and analyzed as ordinal variables.

**Total red meat consumption:** A total red meat consumption variable was created by summing the frequencies for beef (field ID 1369), pork (field ID 1389), and lamb/mutton (field ID 1379) and coded into 4 categories of red meat consumption: <1 time per week, 1.0–1.9 times per week, 2.0–2.9 times per week, and ≥3.0 times per week.[7]

**Total fruit consumption:** One piece of fresh fruit (field ID 1309) and two 'pieces' of dried fruit (field ID 1319) were counted as a serving. The number of total servings consumed per day was summed and grouped into: <2.0 servings per day, 2.0–2.9 servings per day, 3.0–3.9 servings per day, and ≥4.0 servings per day.

**Total vegetable consumption:** Two heaped tablespoons of cooked vegetables (field ID 1289) and salad/raw vegetables (field ID 1299) consumed per day were counted as a serving. The number of total servings consumed per day was summed and grouped into: <2.0 servings per day, 2.0–2.9 servings per day, 3.0–3.9 servings per day, and ≥4.0 servings per day.

**Occupational physical activity:** Responses on whether participants' work involves heavy manual or physical work (field ID 816) or involves mainly walking or standing (field ID 806) were used to create a summary occupational physical activity (OPA) score. According to previous studies,[7,74] the hours of employment per week (field ID 767) were multiplied by 60 (to get the total minutes per week). Values of minutes of employment per week were set to 0 for all participants who did not indicate that they were in paid employment or self-employed (field ID 6142). Then, the number of minutes spent weekly in each type of heavy manual and walking/standing work were calculated by adjusting the total minutes of work per week according to participants' responses. The minutes per week for both types of work were then multiplied by the metabolic equivalent of task (MET) for each activity and these values were summed across both variables to get a total OPA MET value for each participant. In this way, the approximate minutes per week that each participant spent in both heavy manual and walking/standing work were calculated.

**Total sedentary time:** Total sedentary time was measured using self-reported hours spent on a typical day watching television (field ID 1070), using the computer (field ID 1080), and driving (field ID 1090).[75] Values for each variable greater than 24 hours per day were excluded, and those reporting over 16 hours were recoded to 16 hours. Tertiles were used to categorize sedentary time into low (0–4 hours), medium (5–6 hours), and high (>6 hours) levels of sedentary behavior, with 0–4 hours set as the reference and the variable was classed as an unordered factor.

**Body fat composition:** Abdominal and body adipose tissue assessment was performed on the same 1.5T scanner as CMR using a dual-echo Dixon Vibe imaging protocol, generating a dataset with water and fat components separately, facilitating the analysis of body fat composition.[76] Briefly, six overlapping sections were acquired that underwent calibration, stacking, fusion, and segmentation. Body composition analyses were carried out using AMRA Researcher (AMRA Medical AB, Linköping, Sweden). Values for android and gynoid adipose tissue mass were acquired from over 20,000 subjects in the UKB dataset with multi-sequence magnetic resonance and dual-energy X-ray absorptiometry (DXA) scans, using self-supervised, multi-modal alignment for whole-body medical imaging, and transferring segmentation maps from DXA to magnetic resonance imaging scans achieving high accuracy in matching different modality scans without requiring ground-truth magnetic resonance examples.[77]

We also utilized datasets for liver proton density fat fraction employing the gradient echo protocol from the UKB, specifically the Liver MultiScan Dixon method. The LMS Dixon proton density fat fraction captures images during a single breath-hold, reducing motion artifacts and ensuring consistent measurements. Analysis involves three 15-mm circular regions of interest in the liver parenchyma, calculating proton density fat fraction, liver iron concentration, and iron-corrected T1. Proton density fat fraction, a reliable measure of liver fat, is determined using water-fat separation masks, with values over 5% indicating fatty liver disease.[78]

#### Statistical modeling

**Linear models:** Linear models were performed using the R package `stats`.[79] Given $\mathbf{X} = \{X_1, X_2, \ldots, X_N\}$ as the list of covariates, models were fitted with the following equation:

$$Y = \beta_0 + \beta_1 X_1 + \beta_2 X_2 + \cdots + \beta_N X_N + \varepsilon,$$

where $Y$ denotes the outcome variable and $\varepsilon$ represents the error term.

To evaluate whether the effect of a given covariate differs across levels of another covariate, we introduced an interaction term into the linear model. For example, the interaction between $X_1$ and $X_2$ was modeled as:

$$Y = \beta_0 + \beta_1 X_1 + \beta_2 X_2 + \beta_3 (X_1 \times X_2) + \varepsilon,$$

where $\beta_3$ captures the effect modification of $X_1$ by $X_2$.

Parameters were estimated using ordinary least squares by minimizing the sum of squared residuals:

$$\hat{\beta} = \arg\min_{\beta} (Y - X\beta)^\top (Y - X\beta),$$

where $Y$ denotes the outcome vector and $X$ the design matrix. When an interaction term is included, the corresponding column is added to the design matrix $X$, and its coefficient is estimated jointly with all other model parameters using the same least-squares criterion.

**Cox proportional hazards models:** Cox proportional hazards models were fitted using the `survival` package in R.[79] In this semi-parametric model, the hazard function for subject $i$ at time $t$ is expressed as

$$h(t \mid X_i) = h_0(t) \exp\left(\beta^\top X_i\right),$$

where $h_0(t)$ is an unspecified baseline hazard function, $\beta$ denotes the vector of regression coefficients, and $\exp(\beta^\top X_i)$ represents the relative hazard associated with covariates $X_i$.

The coefficients $\beta$ were estimated by maximizing the Cox partial log-likelihood:

$$\ell(\beta) = \sum_{i:\, \delta_i = 1} \left[ \beta^\top X_i - \log\left( \sum_{j \in R(t_i)} \exp\left(\beta^\top X_j\right) \right) \right],$$

where $\delta_i = 1$ indicates an observed event at time $t_i$, $X_i$ denotes the covariate vector for subject $i$, and $R(t_i) = \{j : T_j \geq t_i\}$ is the risk set, defined as the set of subjects still under observation and at risk immediately prior to time $t_i$. Under this definition, right-censored individuals contribute to the partial likelihood up to their censoring time and are removed from the risk set thereafter. The baseline hazard function $h_0(t)$ does not need to be specified for the estimation of $\beta$, rendering the Cox model semi-parametric and allowing inference based solely on the partial likelihood.

**Cox proportional hazards models with restricted cubic splines:** Cox proportional hazards models with restricted cubic splines were fitted using the `survival` and `splines` packages in R.[79] Restricted cubic splines were used to model potential non-linear associations between continuous covariates and the log-hazard.

For a given continuous variable $Z_i$, the spline-augmented predictor is expressed as

$$\eta(Z_i) = \beta_1 Z_i + \sum_{m=1}^{M} \gamma_m B_m(Z_i),$$

where $\beta_1$ denotes the linear effect of $Z_i$, $B_m(\cdot)$ are the restricted cubic spline basis functions determined by the selected knot locations, and $\gamma_m$ are the corresponding spline coefficients.

The linear predictor enters the Cox model as

$$h(t \mid X_i, Z_i) = h_0(t) \exp\left(\beta^\top X_i + \eta(Z_i)\right),$$

where $h_0(t)$ denotes the unspecified baseline hazard and $X_i$ represents the remaining covariates. Coefficients were estimated via maximization of the Cox partial log-likelihood:

$$\ell(\theta) = \sum_{i:\, \delta_i = 1} \left[ \theta^\top W_i - \log\left( \sum_{j \in R(t_i)} \exp\left(\theta^\top W_j\right) \right) \right],$$

where $\theta = (\beta^\top, \gamma_1, \ldots, \gamma_M)^\top$ is the full coefficient vector, $\delta_i = 1$ indicates an observed event, and $W_i$ contains both the standard covariates $X_i$ and the spline-transformed terms $B_m(Z_i)$.

**Fine–Gray subdistribution hazard models:** Fine–Gray subdistribution hazard models were fitted using the *riskRegression* package in R.[79] This approach models covariate effects on the subdistribution hazard for a specific cause of failure in the presence of competing risks, thereby inducing a model for the cumulative incidence function. The subdistribution hazard is specified as

$$\tilde{h}_k(t \mid X_i) = \tilde{h}_{k0}(t)\ \exp\big(\beta^\top X_i\big)\ ,$$

where $\tilde{h}_{k0}(t)$ is an unspecified baseline subdistribution hazard for cause $k$, and $\beta$ denotes the vector of regression coefficients associated with covariates $X_i$.

The Fine–Gray routine estimates $\beta$ by maximizing a weighted partial likelihood for the subdistribution hazard:

$$\ell_{\text{FG}}(\beta) = \sum_{i:\,\delta_i=k} \left[\beta^\top X_i - \log\left(\sum_{j\in\tilde{R}(t_i)} \exp\big(\beta^\top X_j\big)\right)\right],$$

where $\delta_i = k$ indicates that subject $i$ experienced the event of interest, and $\tilde{R}(t_i)$ represents the modified risk set, which includes all individuals who are event-free at $t_i$ and subjects who experienced competing events but are retained according to the Fine–Gray scheme. As in the Cox model, the baseline subdistribution hazard $\tilde{h}_{k0}(t)$ is not required for estimation of $\beta$.

**Weibull accelerated failure time models:** Weibull accelerated failure time models were fitted using the *survival* package in R.[79] In this parametric formulation, the logarithm of survival time is modeled as

$$\log(t_i) = \beta_0 + \beta^\top X_i + \sigma W_i,$$

where $W_i$ follows a standard Gumbel distribution and $\sigma$ is the scale parameter. Under this specification, the survival time $t_i$ follows a Weibull distribution with scale parameter $\lambda_i = \exp(\beta_0 + \beta^\top X_i)$ and shape parameter $k = 1/\sigma$.

The Weibull probability density function is

$$f(t_i \mid \lambda_i, k) = \frac{k}{\lambda_i}\left(\frac{t_i}{\lambda_i}\right)^{k-1} \exp\left[-\left(\frac{t_i}{\lambda_i}\right)^k\right], \qquad t_i \geq 0.$$

Parameters $(\beta, \sigma)$ were estimated by maximizing the full log-likelihood for right-censored survival data:

$$\ell(\beta, \sigma) = \sum_{i=1}^{n} \left\{\delta_i \left[\log k - \log \lambda_i + (k-1)\log\left(\frac{t_i}{\lambda_i}\right)\right] - \left(\frac{t_i}{\lambda_i}\right)^k\right\},$$

where $\delta_i = 1$ denotes an observed event and $\delta_i = 0$ denotes a censored observation.

**Causal mediation analyses:** Causal mediation analyses were performed using the *mediation* package in R.[80] Let $A_i$ denote the exposure (i.e., GlycA), $M_i$ the mediator (i.e., inflammatory protein level), $Y_i$ the outcome, and $X_i$ a vector of baseline covariates for subject $i$.

For continuous outcomes, we specified the following parametric models. The mediator model was given by:

$$M_i = \alpha_0 + \alpha_1 A_i + \alpha_X^\top X_i + \varepsilon_i,$$

and the outcome model was defined as

$$Y_i = \beta_0 + \beta_1 A_i + \beta_2 M_i + \beta_X^\top X_i + \eta_i,$$

where $\varepsilon_i$ and $\eta_i$ are mean-zero error terms.

For time-to-event outcomes, survival time was modeled using the Weibull accelerated failure time specification:

$$\log(T_i) = \beta_0 + \beta_1 A_i + \beta_2 M_i + \beta_X^\top X_i + \sigma W_i,$$

where $T_i$ is the event time, $W_i$ follows a standard Gumbel distribution, and $\sigma$ is the scale parameter.

Under the standard sequential ignorability assumptions, the ACME and average direct effect (ADE) for a contrast between two exposure levels $a$ and $a^*$ were defined in the potential-outcomes framework as

$$\text{ACME}(a) = \mathbb{E}\big[Y_i(a, M_i(a)) - Y_i(a, M_i(a^*))\big],$$

and

$$\text{ADE}(a) = \mathbb{E}\big[Y_i(a, M_i(a^*)) - Y_i(a^*, M_i(a^*))\big],$$

with the total effect decomposed as

$$\mathrm{TE}(a, a^*) = \mathrm{ACME}(a) + \mathrm{ADE}(a).$$

#### Statistical analysis

The relationship between GlycA and CMR-derived IDPs and ECG was examined using multiple linear regression models adjusted for potential confounders. Each model considered each IDP as predicted variable, log-transformed and scaled GlycA as predictor, and was adjusted for BSA, sex, age (at the date of CMR), age$^2$, age:sex interaction and prevalent diabetes (at the time of CMR). The analysis was performed in the pooled cohort, and in separate cohorts according to sex, diabetes, obesity (defined as body mass index >$30 kg/m^2$), and age cohort (grouped as age at CMR < 55, ≤ 56 and < 70, or ≥ 70 years) status. We then calculated the Pearson $r$ correlation between regression coefficients for the association between GlycA and IDPs derived separately in women vs men, obese vs non-obese and diabetic vs non-diabetic. Considering that hsCRP currently represents the most extensively validated inflammatory biomarker,[4] an analogous analysis was conducted substituting log-transformed and scaled hsCRP for GlycA, and Pearson $r$ correlation between regression coefficients was also calculated.

Multivariable linear regressions were also used to explore the association between 80 inflammatory proteins and GlycA. Linear models were built considering GlycA as the outcome variable and each protein as predictor (one protein per model), and adjusted for BSA, sex, age (at the date of CMR), age$^2$, age:sex interaction and prevalent diabetes.

Afterward, a mediation analysis was performed. Each model considered log-transformed and scaled GlycA as exposure variable, each protein as mediator, each IDP as outcome variable, and was adjusted for BSA, sex, age (at the time of recruitment), age$^2$, age:sex interaction and prevalent diabetes. The average causal mediation effect and the proportion of the mediated effect were calculated and are reported with the corresponding 95% CI. An analogous analysis was conducted substituting log-transformed and scaled hsCRP for GlycA.

For the survival analysis, the study cohort was stratified according to quintiles of log-transformed and scaled GlycA, with quintile 1 including the lowest biomarker levels and quintile 5 the highest levels. Each analysis was performed considering GlycA as discrete variable, with quintile 1 as reference level, and also as continuous variable. Results are reported as HR (95% CI).

We employed Cox proportional hazards models to test the hypothesis of GlycA being an independent predictor of the primary and secondary endpoints on top of CMR features or lipid profile after adjusting for confounding factors. Different models were built to progressively correct for confounders: 1) adjusted for BSA, sex, age, age$^2$, age:sex interaction, and statin use; 2) adjusted for BSA, sex, age, age$^2$, age:sex interaction, statin use, and non-cardiovascular death (as a dichotomous confounder); 3) adjusted for BSA, sex, age, age$^2$, age:sex interaction, statin use, low-density lipoprotein (LDL) cholesterol, triglyceride, high-density lipoprotein (HDL) cholesterol, and Lp(a); 4) adjusted for BSA, sex, age, age$^2$, age:sex interaction, statin use, and 12 IDPs derived with least absolute shrinkage and selection operator (LASSO) regression as described below. For models 1), 2), and 3), participants were followed from the time of enrollment until the first occurrence of MACE or death. For model 4), participants were followed from the time of CMR until MACE or death, depending on the specific research question being addressed.

A sensitivity analysis was performed using Fine–Gray subdistribution hazards regression models considering non-cardiovascular death as a competing risk. Models were adjusted for BSA, sex, age, age$^2$, age:sex interaction, statin use, LDL cholesterol, triglyceride, HDL cholesterol, and Lp(a). A further model evaluated the potential role of inflammation on the association between GlycA and events by additionally adjusting for hsCRP.

Non-linear associations between the biomarker and the primary endpoint were explored using Cox proportional hazards models with restricted cubic splines. Restricted cubic spline functions with three knots placed at the 10th, 50th, and 90th percentiles of the biomarker distribution were employed, and the model was adjusted for BSA, sex, age, age$^2$, age:sex interaction, statin use, LDL cholesterol, triglyceride, HDL cholesterol, and Lp(a). The median biomarker value was used as the reference (HR = 1.0). Overall and nonlinearity $P$-values were assessed using likelihood ratio and Wald tests comparing spline and linear terms.

We also performed a mediation analysis considering each protein as a potential mediator of the association between GlycA and incident MACE. For this analysis, we employed a parametric survival model for the outcome using the Weibull distribution and adjusted for confounders and lipid profile. An analogous analysis was conducted substituting log-transformed and scaled hsCRP for GlycA.

We further examined whether interactions between environmental factors and each PRS influence GlycA levels and incident MACE. Multiple linear regression models were adopted to examine whether interactions between environmental factors and each PRS significantly affect GlycA levels. Models were fitted with log-transformed GlycA as the dependent variable and the exposures significantly associated with GlycA (in the pooled, male, and female cohorts) from the previous XWAS as independent variables. All continuous phenotype exposures were scaled and centered to the mean before running the XWAS. For each exposure, all PRS were tested in separate models adding a multiplicative term, and adjusting for BSA, sex, age, age$^2$, age:sex interaction, and ethnicity.

We additionally evaluated whether interactions between PRS and environmental factors modified the association between each exposure and incident MACE, using Cox proportional hazards models and adding a multiplicative term. Models were adjusted for BSA, sex, age, $age^2$, age:sex interaction, and statin use. Patients who experienced MACE before enrollment were excluded. Both analyses were repeated adjusting also for Townsend deprivation index at recruitment (field ID 22189).

Statistical significance was defined as $P < 0.05$ (two-sided). The Benjamini–Hochberg method was applied to control the false discovery rate by adjusting the *P*-values for multiple comparisons.[26]

#### CMR feature selection

Feature selection was performed using LASSO, a penalized regression method that reduces the number of variables. The analysis was performed with the `glmnet` R package.[81] The regularization parameter $\lambda$ controls the strength of the shrinkage to achieve the best compromise between prediction performance and interpretability. The optimal $\lambda$ ($6.11 \times 10^{-4}$) was identified using 10-fold cross-validation and the one standard-error rule, for which the area under the curve (AUC) of the receiver operating characteristic (ROC) of the most parsimonious model is no more than one standard error above the AUC of the ROC of the best model (Supplementary Figure 6).

A total of 14 IDPs were selected: LV ESVi, indexed LV mass, LV global wall thickness, RV EDVi, RV ESVi, RV EF, indexed left atrium minimum volume, left atrium EF, indexed right atrium stroke volume, ascending aorta minimum area, descending aorta distensibility, global circumferential strain, circumferential PDSR, and longitudinal PDSR.

### Supplementary Tables

| Gene | Protein name | Gene | Protein name |
|---|---|---|---|
| **TGF Pathway** | | | |
| tgfb1 | TGF beta-1 proprotein | tgfb2 | TGF beta-2 proprotein |
| tgfbr1 | TGF beta receptor type-1 | tgfbr2 | TGF beta receptor type-2 |
| tgfbr3 | TGF beta receptor type 3 | tgfbi | TGF beta-induced protein ig-h3 |
| **TNF Family** | | | |
| tnf | TNF | tnfsf8 | TNF ligand superfamily member 8 |
| tnfsf10 | TNF ligand superfamily member 10 | tnfsf11 | TNF ligand superfamily member 11 |
| tnfsf12 | TNF ligand superfamily member 12 | tnfsf13 | TNF ligand superfamily member 13 |
| tnfsf13b | TNF ligand superfamily member 13B | tnfsf14 | TNF ligand superfamily member 14 |
| tnfrsf1a | TNF receptor superfamily member 1A | tnfrsf1b | TNF receptor superfamily member 1B |
| ltbr | TNF receptor superfamily member 3 | tnfrsf4 | TNF receptor superfamily member 4 |
| tnfrsf6b | TNF receptor superfamily member 6B | tnfrsf8 | TNF receptor superfamily member 8 |
| tnfrsf9 | TNF receptor superfamily member 9 | tnfrsf10a | TNF receptor superfamily member 10A |
| tnfrsf10b | TNF receptor superfamily member 10B | tnfrsf10c | TNF receptor superfamily member 10C |
| tnfrsf11a | TNF receptor superfamily member 11A | tnfrsf11b | TNF receptor superfamily member 11B |
| tnfrsf12a | TNF receptor superfamily member 12A | tnfrsf13b | TNF receptor superfamily member 13B |
| tnfrsf13c | TNF receptor superfamily member 13C | tnfrsf14 | TNF receptor superfamily member 14 |
| tnfrsf17 | TNF receptor superfamily member 17 | tnfrsf19 | TNF receptor superfamily member 19 |
| tnfrsf21 | TNF receptor superfamily member 21 | tnfaip2 | TNF alpha-induced protein 2 |
| tnfaip8 | TNF alpha-induced protein 8 | tnfaip8l2 | TNF alpha-induced protein 8-like 2 |
| **Interleukins and Receptors** | | | |
| il1r1 | IL-1 receptor type 1 | il1r2 | IL-1 receptor type 2 |
| il1rl1 | IL-1 receptor-like 1 | il1rl2 | IL-1 receptor-like 2 |
| il1rap | IL-1 receptor accessory protein | il1rn | IL-1 receptor antagonist protein |
| il2ra | IL-2 receptor subunit alpha | il4r | IL-4 receptor subunit alpha |
| il6 | IL-6 | il10 | IL-10 |
| il10rb | IL-10 receptor subunit beta | il12b | IL-12 subunit beta |
| il17ra | IL-17 receptor A | il18 | IL-18 |
| il18r1 | IL-18 receptor 1 | il1b | IL-1 beta |
| osm | Oncostatin-M | | |
| **Chemokines** | | | |
| ccl3 | C-C motif chemokine 3 | ccl4 | C-C motif chemokine 4 |
| ccl11 | Eotaxin | ccl19 | C-C motif chemokine 19 |
| ccl20 | C-C motif chemokine 20 | cxcl1 | Growth-regulated alpha protein |
| cxcl8 | IL-8 | cxcl10 | C-X-C motif chemokine 10 |
| **Matrix Metalloproteinases** | | | |
| mmp1 | Interstitial collagenase | mmp3 | Stromelysin-1 |
| mmp7 | Matrilysin | mmp8 | Neutrophil collagenase |
| mmp9 | Matrix metalloproteinase-9 | mmp10 | Stromelysin-2 |
| mmp12 | Macrophage metalloelastase | mmp13 | Collagenase 3 |
| mmp15 | Matrix metalloproteinase-15 | | |
| **Cell Surface Receptors and Adhesion Molecules** | | | |
| cd4 | T-cell surface glycoprotein CD4 | itgb2 | Integrin beta-2 |
| cd40 | TNF receptor superfamily member 5 | cd40lg | CD40 ligand |
| pdlim7 | PDZ and LIM domain protein 7 | | |
| **Growth Factors** | | | |
| hgf | Hepatocyte growth factor | csf1 | Macrophage colony-stimulating factor 1 |
| **Signaling Proteins and Kinases** | | | |
| src | Proto-oncogene tyrosine-protein kinase Src | casp8 | Caspase-8 |

**Supplementary Table 1.** Data dictionary of proteins from the proteomics dataset provided by UK Biobank. **Abbreviations:** IL, interleukin; TGF, Transforming Growth Factor; TNF, Tumor Necrosis Factor

| Exposure factor | UKB data-field ID(s) |
|---|---|
| *Early life factors* | |
| Country of birth, Breastfed, Body size at age 10, Height at age 10, Handedness, Part of multiple birth, Maternal smoking, Birth weight | 1647, 1677, 1687, 1697, 1707, 1777, 1787, 20022 |
| *Social support* | |
| Frequency of family visits, Frequency of confiding in someone, Loneliness, Adult education, Pub attendance, Religious group attendance | 1031, 2110, 2020, 6163, 6161, 6162 |
| *Household* | |
| Accommodation type, Own or rent, Years at address, Household size, Household income, Household partner, Household children, Household grandchild, Gas fire heating, Gas hob heating, Open fire heating, Gas central heating, Electric storage heating, Oil central heating | 670, 680, 699, 709, 738, 6141, 6141, 6141, 6139, 6139, 6139 |
| *Mental health* | |
| Neuroticism score, Mood swings, Miserableness, Irritability, Sensitivity, Fed-up feelings, Nervous feelings, Worrier, Tense, Worry after embarrassment, Nerves, Guilty feelings, Risk taking, Depressed mood, Unenthusiasm, Tenseness, Tiredness | 20127, 1920, 1930, 1940, 1950, 1960, 1970, 1980, 1990, 2000, 2010, 2030, 2040, 2050, 2060, 2070, 2080 |
| *Electronics use* | |
| Weekly mobile phone use, Use of speakerphone, Computer games | 1120, 1130, 2237 |
| *Sleep* | |
| Ease of waking, Chronotype, Daytime napping, Sleep difficulties, Snoring | 1170, 1180, 1190, 1200, 1210 |
| Sleep duration | derived |
| *Smoking* | |
| Ever smoked, Pack years proportion, Pack years, Smoking status, Tobacco use | 20160, 20162, 20161, 20116, 1239 |
| *Alcohol* | |
| Alcohol drinking frequency | 1558 |
| *Diet* | |
| Oily fish, Non-oily fish, Processed meat, Poultry, Cheese, Salt added to food, Tea, Coffee, Water, Diet change in last 5 years, Diet variation | 1329, 1339, 1349, 1359, 1408, 1478, 1488, 1498, 1528, 1538, 1548 |
| Bread fibre, Cereal fibre, Total fruit, Total vegetables, Red meat | derived |
| *Sun exposure* | |
| Summer outdoors time, Winter outdoors time, Ease of tanning, Childhood sunburn episodes, Sun protection use, Solarium use | 1050, 1060, 1727, 1737, 2267, 2277 |
| *Sexual history* | |
| Same-sex intercourse | 2159 |
| *Physical activity* | |
| IPAQ activity group, Leisure walks (past 4 weeks), Heavy DIY (past 4 weeks), Light DIY (past 4 weeks), Other exercise (past 4 weeks), Strenuous sports (past 4 weeks), Gym attendance | 22032, 6164, 6164, 6164, 6164, 6164, 6160 |
| Occupational physical activity (OPA), Sedentary behaviour | derived |
| *Material deprivation* | |
| Townsend deprivation index | 189 |
| *Physical environment* | |
| Proximity to major road, Inverse distance to nearest major road, Inverse distance to nearest road, Nitrogen dioxide (2005–2010), Nitric oxide (2010), $PM_{10}$ (2007–2010), $PM_{2.5}$ absorbance (2010), $PM_{2.5}$ (2010), $PM_{2.5-10}$ (2010), Average daytime noise, Average evening noise, Average night noise, Natural environment buffer (300 m), Natural environment buffer (1000 m), Greenspace buffer (300 m), Greenspace buffer (1000 m), Domestic garden buffer (300 m), Domestic garden buffer (1000 m), Water buffer (300 m), Water buffer (1000 m), Distance to coast, Population density, Road length within 100 m, Traffic load on major roads, Traffic intensity on nearest major road, Traffic intensity on nearest road | 24014, 24012, 24010, 24016, 24017, 24018, 24003, 24004, 24019, 24005, 24007, 24006, 24008, 24023, 24024, 24020, 24021, 24022, 24506, 24507, 24500, 24503, 24501, 24504, 24502, 24505, 24508, 20118, 24015, 24013, 24011, 24009 |
| *Stressful life events* | |
| Death of a relative, Death of a partner, Financial difficulties, Divorce | 6147, 6148, 6150, 6149 |
| *Employment* | |
| Volunteering, Student status, Employment status, Homemaker, Retired, Unemployed | 6160, 6142, 6142, 6142, 6142, 6142 |
| *Supplements* | |
| Calcium, Fish oil, Glucosamine, Iron, Selenium, Zinc, Folate, Multivitamins, Vitamin A, Vitamin B, Vitamin C, Vitamin D, Vitamin E | 6179, 6179, 6179, 6179, 6179, 6179, 6155, 6155, 6155, 6155, 6155, 6155, 6155 |

**Supplementary Table 2.** Data dictionary of exposure variables with UK Biobank field IDs.

| Body fat composition | Field ID | Count |
|---|---|---|
| Visceral adipose tissue | 23288 | 40,021 |
| Abdominal subcutaneous adipose tissue | 22408 | 55,939 |
| Muscle adipose tissue infiltration | 22435 | 39,319 |
| Liver proton density fat fraction | 22436 | 9,879 |
| Android fat mass | 23245 | 70,528 |
| Gynoid fat mass | 23262 | 70,528 |
| Trunk fat mass | 23284 | 70,528 |
| Total fat mass | 23278 | 70,528 |

**Supplementary Table 3.** Body fat composition measures with corresponding UK Biobank field IDs and total count.

| Diagnostic label | First Occur. ICD-10 | ICD-10 | ICD-9 | Self-report | OPSC-4 |
|---|---|---|---|---|---|
| *Cardiovascular risk factors* | | | | | |
| Hypertension | - | I100, I110, I119, I120, I129-I132, I139, I150-I152, I159 | 4010, 4011, 4019, 4039, 404, 4040, 4041, 4049, 405, 4050, 4051, 4059 | 1065, 1072, 1073 | - |
| Diabetes | - | E100-E119, E121, E123, E125, E128-E149 | 250, 2500, 25001, 25009, 2501, 25010, 25011, 25019, 2502, 25020, 25021, 25029, 2503-2507, 2509, 25090, 25091, 25099 | 2443, 1220-1223, 1276, 1468, 1607 | - |
| Hypercholesterolaemia | - | E780, E782 | 2720, 27202, 27209, 27200, 2720 | 1473 | - |
| Obesity | - | E66 | 278, 2780, 27800, 27801 | - | - |
| General CVD | - | I00, I010, I011, I018, I019, I020, I029, I050-I052, I058-I062, I068-I072, I078-I083, I088-I091, I098, I099, I10, I110, I119, I120, I129, I130-I132, I139, I150-I152, I158, I159, I200, I201, I208, I209, I21, I210-I214, I219, I22, I220, I221, I228, I229, I23, I230-I236, I238, I24, I240, I241, I248, I249, I25, I250-I253, I254-I256, I258, I259, I30, I300, I301, I308, I309, I31, I310-I313, I318, I319, I32, I320, I321, I328, I33, I330, I339, I34, I340-I342, I348, I349, I35, I350-I352 | 390-398, 401-405, 410-417, 420-429, 430-438, 440-448, 451-459 | 1473 | - |
| *Outcomes* | | | | | |
| Angina | I20 | I200, I201, I208, I209 | 4139 | 1074 | - |
| MI | I21*, I22*, I23* | I21, I210-I214, I219, I22, I220, I221, I228, I229, I23, I230-I236, I238, I24, I248, I249, I252 | 410, 4109, 411, 4119, 412, 4129 | 1471, 1483 | - |
| IHD | I20*, I21*, I22*, I23*, I24*, I25* | I20, I200, I201, I208, I209, I21, I210-I214, I219, I21X, I22, I220, I221, I228, I229, I23, I230-I236, I238, I24, I240, I241, I248, I249, I25, I250-I256, I258, I259 | 410, 4109, 411, 4119, 412, 4129, 413, 4139, 414, 4140, 4141, 4148, 4149 | 1074, 1075 | 1523, 1070, 1071, 1095 |
| CKD | N18*, N19* | N18, N181-N185, N188, N189, Y841, Z49, Z490-Z492, Z992 | 585, 5859, 586, 5869, V541, V56, V560, V568 | 1192-1194 | M01, M026, M027, M084, M17, L746 |
| AF | I48* | I48, I480-I484, I489 | 4273 | 1471, 1483 | - |
| Cardiac Arrest | I46* | I46, I460, I461, I469, I490 | 4274, 4275 | - | - |
| Heart Failure | I50* | I110, I130, I132, I500, I501, I509 | 428, 4280, 4281, 4289 | 1076 | - |
| Heart Transplant | - | - | - | - | K01, K02 |
| Stroke/CVE | I60*, I61*, I62*, I63*, I64* | I60, I600-I609, I61, I610-I616, I618-I621, I62, I629, I63, I630-I636, I638, I639, I64 | 430, 4309, 431, 4319, 432, 4320, 4321, 4329, 433, 4330-4333, 4338, 4339, 434, 4340, 4341, 4349 | 1081, 1583, 1083, 1086 | - |

**Supplementary Table 4.** Diagnostic labels and associated coding systems in UK Biobank. ICD: International Classification of Diseases; Self-report: self-reported outcome; OPSC-4: Office of Population Censuses and Surveys Classification of Interventions and Procedures, version 4. **Abbreviations:** AF, atrial fibrillation; CKD, chronic kidney disease; CVD, cardiovascular disease; CVE, cerebrovascular events; IHD, ischaemic heart disease; MI, myocardial infarction.

| **Polygenic risk score** (n=485,717) | **Field ID** |
|---|---|
| Cardiovascular disease | 26223 |
| Resting heart rate | 21150 |
| Total cholesterol | 21151 |
| HLD cholesterol | 26242 |
| LDL cholesterol | 26250 |
| Total triglyceride | 21152 |
| Type 2 diabetes | 26285 |
| Coronary artery disease | 26227 |
| Arterial hypertension | 26244 |
| Stroke | 26248 |

**Supplementary Table 5.** Polygenic risk scores for cardiovascular disease available for 485,717 individuals in the UK Biobank with corresponding field IDs. **Abbreviations:** HDL, high-density lipoprotein; LDL, low-density lipoprotein.

| Characteristic | Overall (n = 70,809) | Quintile 1 (n = 17,988) | Quintile 2 (n = 15,688) | Quintile 3 (n = 14,262) | Quintile 4 (n = 12,554) | Quintile 5 (n = 10,316) | p-value |
|---|---|---|---|---|---|---|---|
| **Demographics** | | | | | | | |
| Age at MRI, years | 66.2 ± 8.0 | 64.9 ± 8.14 | 66.2 ± 8.0 | 66.7 ± 7.9 | 67.0 ± 7.8 | 66.9 ± 7.8 | <0.001 |
| BMI, kg/$m^2$ | 26.62 ± 4.44 | 24.91 ± 3.79 | 26.08 ± 4.08 | 26.97 ± 4.32 | 27.76 ± 4.47 | 28.54 ± 4.88 | <0.001 |
| BSA, $m^2$ | 1.88 ± 0.22 | 1.82 ± 0.21 | 1.86 ± 0.22 | 1.89 ± 0.22 | 1.92 ± 0.22 | 1.94 ± 0.22 | <0.001 |
| DBP, mmHg | 79.0 ± 10.2 | 77.5 ± 10.2 | 78.8 ± 10.1 | 79.3 ± 10.1 | 79.9 ± 10.1 | 80.3 ± 10.0 | <0.001 |
| SBP, mmHg | 142.1 ± 19.5 | 138.0 ± 19.7 | 141.5 ± 19.4 | 143.1 ± 19.1 | 144.6 ± 19.2 | 145.6 ± 19.1 | <0.001 |
| MAP, mmHg | 97.1 ± 13.9 | 94.9 ± 14.0 | 96.7 ± 13.8 | 97.6 ± 13.6 | 98.5 ± 13.9 | 99.2 ± 13.9 | <0.001 |
| Glycoprotein acetyls, mmol/L | 0.80 ± 0.12 | 0.66 ± 0.05 | 0.75 ± 0.02 | 0.81 ± 0.02 | 0.88 ± 0.02 | 0.99 ± 0.07 | <0.001 |
| Time to MRI, years | 11.4 ± 3.0 | 11.4 ± 3.0 | 11.3 ± 3.0 | 11.3 ± 3.0 | 11.4 ± 3.0 | 11.4 ± 3.0 | 0.002 |
| **Cardiac magnetic resonance** | | | | | | | |
| LVEF, % | 59.6 ± 6.4 | 59.5 ± 6.2 | 59.6 ± 6.4 | 59.7 ± 6.5 | 59.7 ± 6.6 | 59.7 ± 6.6 | 0.281 |
| LVEDVi, mL/$m^2$ | 77.5 ± 14.4 | 80.8 ± 14.4 | 78.2 ± 14.2 | 76.7 ± 14.1 | 75.6 ± 14.1 | 74.4 ± 14.0 | <0.001 |
| LVESVi, mL/$m^2$ | 30.3 (25.5-36.1) | 31.74 (26.9-37.4) | 30.5 (25.8-36.4) | 29.9 (25.2-35.7) | 29.5 (24.8-35.2) | 29.1 (24.3-34.8) | <0.001 |
| LVSVi, mL/$m^2$ | 46.0 ± 8.6 | 47.9 ± 8.7 | 46.4 ± 8.5 | 45.5 ± 8.4 | 44.9 ± 8.4 | 44.1 ± 8.2 | <0.001 |
| LVMi, g/$m^2$ | 45.2 ± 8.5 | 45.1 ± 8.4 | 45.2 ± 8.5 | 45.2 ± 8.5 | 45.3 ± 8.6 | 45.2 ± 8.5 | 0.644 |
| LVCO, L/min | 5.4 ± 1.3 | 5.3 ± 1.2 | 5.4 ± 1.2 | 5.4 ± 1.3 | 5.5 ± 1.3 | 5.5 ± 1.3 | <0.001 |
| LVCi, L/min/$m^2$ | 2.9 ± 0.6 | 2.9 ± 0.6 | 2.9 ± 0.6 | 2.9 ± 0.6 | 2.9 ± 0.6 | 2.9 ± 0.6 | <0.001 |
| RVEF, % | 57.2 ± 6.5 | 57.0 ± 6.3 | 57.2 ± 6.5 | 57.3 ± 6.6 | 57.3 ± 6.6 | 57.5 ± 6.5 | <0.001 |
| RVEDVi, mL/$m^2$ | 82.3 ± 15.4 | 86.0 ± 15.6 | 83.1 ± 15.4 | 81.4 ± 15.0 | 80.0 ± 14.9 | 78.3 ± 14.8 | <0.001 |
| RVESVi, mL/$m^2$ | 35.4 ± 9.4 | 37.2 ± 9.7 | 35.8 ± 9.5 | 34.0 ± 9.2 | 34.3 ± 9.1 | 33.5 ± 9.0) | <0.001 |
| RVSVi, mL/$m^2$ | 46.9 ± 9.1 | 48.8 ± 9.1 | 47.3 ± 9.1 | 46.4 ± 9.0 | 45.6 ± 8.9 | 44.8 ± 8.7 | <0.001 |
| WT Global, mm | 5.7 ± 0.8 | 5.53 ± 0.7 | 5.7 ± 0.8 | 5.75 ± 0.8 | 5.8 ± 0.8 | 5.9 ± 0.8 | <0.001 |
| WT Max Global, mm | 9.4 (8.4-10.5) | 9.0 (8.0-10.1) | 9.3 (8.3-10.4) | 9.4 (8.5-10.6) | 9.6 (8.6-10.7) | 9.7 (8.7-10.8) | <0.001 |
| Ecc global, abs(%) | 22.3 ± 3.6 | 22.4 ± 3.4 | 22.3 ± 3.6 | 22.3 ± 3.6 | 22.2 ± 3.7 | 22.2 ± 3.7 | <0.001 |
| Ell global, abs(%) | 18.5 ± 2.9 | 18.4 ± 2.9 | 18.5 ± 2.9 | 18.5 ± 3.0 | 18.5 ± 3.0 | 18.5 ± 3.0 | 0.004 |
| Err global, abs(%) | 45.7 ± 8.8 | 45.5 ± 8.5 | 45.7 ± 8.8 | 45.8 ± 8.9 | 45.7 ± 9.0 | 45.9 ± 9.0 | <0.001 |
| PDSR longitudinal, abs(%) | 1.6 ± 0.6 | 1.7 ± 0.6 | 1.6 ± 0.6 | 1.6 ± 0.6 | 1.5 ± 0.5 | 1.5 ± 0.5 | <0.001 |
| PDSR circumferential, abs(%) | 2.3 ± 0.7 | 2.4 ± 0.7 | 2.3 ± 0.7 | 2.2 ± 0.7 | 2.1 ± 0.7 | 2.1 ± 0.7 | <0.001 |
| PDSR radial, abs(%) | 5.7 ± 1.9 | 6.1 ± 1.9 | 5.8 ± 1.9 | 5.6 ± 1.8 | 5.5 ± 1.8 | 5.4 ± 1.8 | <0.001 |
| LAEF, % | 60.0 ± 9.7 | 60.0 ± 9.5 | 60.1 ± 9.7 | 60.0 ± 9.7 | 59.8 ± 10.0 | 59.9 ± 9.9 | 0.061 |
| LASVi, mL/$m^2$ | 22.9 ± 5.7 | 23.6 ± 5.9 | 23.0 ± 5.7 | 22.7 ± 5.7 | 22.4 ± 5.6 | 22.2 ± 5.5 | <0.001 |

| | | | | | | | |
|---|---|---|---|---|---|---|---|
| LAVmax, mL/m$^2$ | 38.0 (31.2-45.7) | 39.2 (32.1-46.9) | 38.3 (31.4-46.0) | 37.7 (31.0-45.1) | 37.4 (30.6-44.9) | 37.2 (30.4-44.4) | <0.001 |
| LAVmin, mL/m$^2$ | 14.9 (11.1-19.5) | 15.4 (11.5-20.0) | 15.0 (11.1-19.7) | 14.8 (11.0-20.0) | 14.7 (10.9-19.2) | 14.6 (10.8-19.0) | <0.001 |
| RAEF, % | 46.7 ± 9.1 | 46.3 ± 8.9 | 46.6 ± 9.0 | 46.8 ± 9.1 | 47.0 ± 9.3 | 47.2 ± 9.2 | <0.001 |
| RASVi, mL/m$^2$ | 21.4 ± 6.9 | 23.0 ± 7.1 | 21.8 ± 6.9 | 21.0 ± 6.8 | 20.3 ± 6.7 | 19.6 ± 6.4 | <0.001 |
| RAVmax, mL/m$^2$ | 44.5 (36.7-53.6) | 48.2 (40.2-57.8) | 45.4 (37.7-54.8) | 43.8 (36.1-52.5) | 42.1 (34.9-50.8) | 40.5 (33.4-48.8) | <0.001 |
| RAVmin, mL/m$^2$ | 23.3 (18.4-29.4) | 25.5 (20.4-32.1) | 23.8 (19.0-30.0) | 22.9 (18.1-28.7) | 22.1 (17.5-27.7) | 21.08 (16.6-26.5) | <0.001 |
| AAo max area, mm$^{-2}$ | 869 ± 194 | 854 ± 196 | 867 ± 195 | 873 ± 192 | 878 ± 192 | 880 ± 193.55 | <0.001 |
| AAo min area, mm$^2$ | 794 ± 189 | 778 ± 192 | 792 ± 189 | 800 ± 187 | 804 ± 185 | 806 ± 187 | <0.001 |
| AAo distensibility, $10^{-3}$mmHg$^{-1}$ | 1.3 (0.9-2.0) | 1.4 (1.0-2.2) | 1.3 (0.9-2.1) | 1.3 (0.9-1.9) | 1.3 (0.9-1.9) | 1.3 (0.9-1.9) | <0.001 |
| DAo max area, mm$^2$ | 483 ± 100 | 471 ± 99 | 481 ± 101 | 486 ± 99 | 491 ± 100 | 491 ± 101 | <0.001 |
| DAo min area, mm$^2$ | 429 ± 95 | 416 ± 95 | 427 ± 96 | 433 ± 94 | 437 ± 94 | 437 ± 95 | <0.001 |
| DAo distensibility, $10^{-3}$mmHg$^{-1}$ | 1.9 (1.3-2.6) | 2.1 (1.4-2.9) | 1.9 (1.3-2.7) | 1.8 (1.3-2.6) | 1.8 (1.3-2.5) | 1.8 (1.3-2.4) | <0.001 |
| **Electrocardiogram** | | | | | | | |
| VentricularRate, beats/min | 61 (55-68) | 59 (53-65) | 60 (54-67) | 61 (55-68) | 62 (56-69) | 63 (57-71) | <0.001 |
| PQInterval, ms | 164 (148-182) | 164 (148-182) | 164 (148-182) | 164 (148-182) | 164 (148-182) | 164 (148-182) | 0.024 |
| PDuration, ms | 97 ± 17 | 98 ± 17 | 97 ± 17 | 97 ± 17 | 97 ± 17 | 97 ± 17 | 0.006 |
| QRSDuration, ms | 86 (80-94) | 86 (80-94) | 86 (80-94) | 86 (80-94) | 86 (80-94) | 86 (80-94) | 0.747 |
| QTInterval, ms | 420 ± 32 | 425 ± 32 | 421 ± 32 | 418 ± 31.5 | 414 ± 32 | 415 ± 32 | <0.001 |
| QTCInterval, ms | 421 (406-437) | 419 (404-435) | 420 (406-436) | 422 (406-438) | 423 (407-439) | 424 (409-440) | <0.001 |
| RRInterval, ms | 993 ± 165 | 1,028 ± 167 | 1,003 ± 166 | 986 ± 160 | 971 ± 159 | 956 ± 161 | <0.001 |
| PPInterval, ms | 986 ± 185 | 1,020 ± 187 | 996 ± 187 | 979 ± 181 | 963 ± 182 | 949 ± 177 | <0.001 |
| PAxis, ° | 50 ± 24 | 52 ± 24 | 51 ± 24 | 49 ± 24 | 48.4 ± 23.5 | 47.8 ± 23.7 | <0.001 |
| RAxis, ° | 26 ± 38 | 32 ± 38 | 27 ± 38 | 23 ± 37 | 21 ± 37 | 20 ± 36 | <0.001 |
| TAxis, ° | 40 (23-56) | 44 (27-59) | 41 (23-57) | 39 (21-55) | 38 (20-55) | 37 (20-55) | <0.001 |
| POnset, ms | 271 ± 30 | 272 ± 30 | 271 ± 30 | 271 ± 30 | 270 ± 30 | 271 ± 30 | <0.001 |
| POffset, ms | 372 (352-390) | 374 (354-390) | 372 (352-390) | 372 (350-388) | 370 (350-388) | 370 (350-388) | <0.001 |
| QOnset, ms | 438 (432-444) | 440 (434-444) | 438 (432-444) | 438 (432-444) | 438 (430-444) | 438 (430-444) | <0.001 |
| QOffset, ms | 524 (516-532) | 524 (518-532) | 524 (516-532) | 524 (516-532) | 524 (516-532) | 524 (516-532) | <0.001 |
| TOffset, ms | 857 ± 33 | 863 ± 32 | 858 ± 32 | 856 ± 32 | 853 ± 33 | 851 ± 33 | <0.001 |

**Supplementary Table 6. Cardiac MRI, physiological, and electrocardiographic characteristics across GlycA quintiles.** Values are mean ± SD or median [interquartile range]. **Abbreviations:** AAo, ascending aorta; BSA, body surface area; BMI, body mass index; DAo, descending aorta; DBP, diastolic blood pressure; EDVi, end-diastolic volume indexed; Ell, longitudinal strain; Ecc, circumferential strain; Err, radial strain; ESVi, end-systolic volume indexed; GlycA, glycoprotein acetyl; LA, left atrium; LAEF, left atrial ejection fraction; LASVi, left atrial stroke volume indexed; LAVmax, maximum left atrial volume indexed; LAVmin, minimum left atrial volume indexed; LV, left ventricle; LVCO, left ventricular cardiac output; LVCi, left ventricular cardiac index; LVEDVi, left ventricular end-diastolic volume indexed; LVEF, left ventricular ejection fraction; LVESVi, left ventricular end-systolic volume indexed; LVMi, left ventricular mass indexed; LVSVi, left ventricular stroke volume indexed; MAP, mean arterial pressure; PDSR, peak diastolic strain rate; P, P wave; Q, Q wave; QRS, QRS interval; R, R wave; RA, right atrium; RAEF, right atrial ejection fraction; RASVi, right atrial stroke volume indexed; RAVmax, maximum right atrial volume indexed; RAVmin, minimum right atrial volume indexed; RV, right ventricle; RVEDVi, right ventricular end-diastolic volume indexed; RVEF, right ventricular ejection fraction; RVESVi, right ventricular end-systolic volume indexed; RVSVi, right ventricular stroke volume indexed; SBP, systolic blood pressure; SVi, stroke volume indexed; T, T wave; WT, myocardial wall thickness.

| Endpoint | Full metabolomic cohort<br>N = 488,079 | Previous MACE excluded<br>N = 454,821 | CMR + ECG + Metabolomic<br>N = 72,541 |
|---|---|---|---|
| MACE (incident) | 74,336 (15%) | 58,971 (13%) | 2,034 (2.8%) |
| MACE (prevalent) | 33,258 (6.8%) | - | - |
| Myocardial infarction | 37,450 (7.7%) | 27,246 (6.0%) | 931 (1.3%) |
| Stroke | 32,164 (6.6%) | 27,658 (6.1%) | 933 (1.3%) |
| Heart failure | 21,118 (4.3%) | 14,979 (3.3%) | 391 (0.5%) |
| Cardiovascular death | 10,377 (2.1%) | 7,382 (1.6%) | 180 (0.2%) |
| All-cause death | 55,076 (11%) | 45,642 (10%) | 1,247 (1.7%) |

**Supplementary Table 7. Incident outcomes.** Number of incident outcomes among with available metabolomic data and metabolomic, CMR and ECG. Cardiovascular death is defined as death due to myocardial infarction, stroke or heart failure. **Abbreviations:** CMR, cardiac magnetic resonance; ECG, electrocardiogram; MACE, major adverse cardiovascular events.

| | Hazard ratio (95% confidence interval) | | | | | |
|---|---|---|---|---|---|---|
| **Model** | per SD | Q1 | Q2 | Q3 | Q4 | Q5 |
| Covariate-adjusted | 1.20 (1.19–1.21) | 1 (ref) | 1.13 (1.09–1.16) | 1.21 (1.17–1.24) | 1.34 (1.31–1.38) | 1.64 (1.60–1.69) |
| Covariate + other death | 1.21 (1.20–1.22) | 1 (ref) | 1.13 (1.10–1.16) | 1.21 (1.18–1.25) | 1.36 (1.32–1.40) | 1.67 (1.63–1.72) |
| Covariate + lipid profile | 1.16 (1.15–1.17) | 1 (ref) | 1.08 (1.05–1.12) | 1.13 (1.09–1.17) | 1.25 (1.21–1.29) | 1.46 (1.41–1.52) |
| Covariate + other death + lipid profile (Fine-Gray) | 1.21 (1.19–1.23) | 1 (ref) | 1.09 (1.05–1.12) | 1.12 (1.09–1.17) | 1.24 (1.20–1.28) | 1.43 (1.38–1.49) |
| Covariate + other death + lipid profile + hsCRP (Fine-Gray) | 1.05 (1.04–1.07) | 1 (ref) | 1.03 (0.99–1.06) | 1.02 (0.98–1.06) | 1.07 (1.03–1.11) | 1.14 (1.09–1.18) |
| Covariate + IDPs | 1.13 (1.08–1.19) | 1 (ref) | 1.06 (0.90–1.23) | 1.14 (0.98–1.33) | 1.23 (1.06–1.43) | 1.38 (1.19–1.61) |

**Supplementary Table 8. Hazard ratios for first major cardiovascular events according to standard deviation increase and quintiles of increasing glycoprotein acetyls levels.** Glycoprotein acetyls was log-transformed and scaled. Lipid profile included: total cholesterol, low-density lipoprotein, very low-density lipoprotein, high-density lipoprotein, remnant cholesterol, total triglycerides, Lp(a). Imaging-derived phenotypes (IDPs) included: indexed left ventricle (LV) end-systolic volume, indexed LV mass, LV global wall thickness, right ventricle (RV) end-diastolic volume, RV end-systolic volume, RV ejection fraction (EF), right atrial EF, indexed left atrial minimum volume, left atrial EF, indexed right atrium stroke volume, ascending aorta minimum area, descending aorta distensibility, global circumferential strain, circumferential peak-diastolic strain rate, longitudinal peak-diastolic strain rate. All models were adjusted for body surface area, sex, age, $age^2$, age:sex interaction, statin use. Participants were followed from enrollment until first occurrence of major cardiovascular events (MACE) for the lipid-adjusted model, or from cardiac magnetic resonance until first MACE for the IDPs-adjusted model.

| Model | Hazard ratio (95% CI) | | | | | |
|---|---|---|---|---|---|---|
| | per SD | Q1 | Q2 | Q3 | Q4 | Q5 |
| Covariate-adjusted | 1.25 (1.24–1.26) | 1 (ref) | 1.13 (1.10–1.17) | 1.27 (1.23–1.31) | 1.48 (1.44–1.52) | 1.86 (1.80–1.91) |
| Covariate + other death | 1.26 (1.25–1.27) | 1 (ref) | 1.13 (1.10–1.17) | 1.27 (1.23–1.31) | 1.50 (1.45–1.54) | 1.90 (1.84–1.95) |
| Covariate + lipid profile | 1.22 (1.21–1.23) | 1 (ref) | 1.11 (1.07–1.15) | 1.22 (1.17–1.26) | 1.38 (1.34–1.43) | 1.71 (1.65–1.77) |
| Covariate + other death + lipid profile (Fine-Gray) | 1.20 (1.19–1.21) | 1 (ref) | 1.11 (1.07–1.15) | 1.21 (1.17–1.26) | 1.37 (1.32–1.42) | 1.65 (1.60–1.71) |
| Covariate + IDPs | 1.14 (1.09–1.20) | 1 (ref) | 1.06 (0.90–1.25) | 1.13 (0.97–1.33) | 1.35 (1.16–1.58) | 1.42 (1.21–1.66) |

**Supplementary Table 9. Hazard ratios for first major cardiovascular events according to standard deviation increase and quintiles of increasing high-sensitivity C-reactive protein levels.** high-sensitivity C-reactive protein was log-transformed and scaled. Lipid profile included: total cholesterol, LDL, VLDL, HDL, remnant cholesterol, total triglycerides, Lp(a). Imaging-derived phenotypes (IDPs) included: indexed left ventricle (LV) end-systolic volume, indexed LV mass, LV global wall thickness, right ventricle (RV) end-diastolic volume, RV end-systolic volume, RV ejection fraction (EF), indexed left atrium minimum volume, left atrium EF, indexed right atrium stroke volume, ascending aorta minimum area, descending aorta distensibility, global circumferential strain, circumferential peak-diastolic strain rate, longitudinal peak-diastolic strain rate. All models were adjusted for body surface area, sex, age, age$^2$, age:sex interaction, statin use. Participants were followed from enrollment until first occurrence of major cardiovascular events (MACE) for the lipid-adjusted model, or from the date of the cardiac magnetic resonance imaging until first MACE for the IDPs-adjusted model.

| Model | Hazard ratio (95% CI) per SD | Q1 | Q2 | Q3 | Q4 | Q5 |
|---|---|---|---|---|---|---|
| **Glycoprotein acetyl** | | | | | | |
| Covariate-adjusted | 1.22 (1.22–1.24) | 1 (ref) | 1.05 (1.02–1.09) | 1.16 (1.12–1.19) | 1.28 (1.25–1.32) | 1.67 (1.63–1.72) |
| Covariate + lipid profile | 1.28 (1.27–1.30) | 1 (ref) | 1.09 (1.05–1.13) | 1.22 (1.17–1.26) | 1.41 (1.36–1.47) | 1.85 (1.79–1.93) |
| Covariate + lipid profile + hsCRP | 1.14 (1.12–1.15) | 1 (ref) | 1.01 (1.97–1.05) | 1.06 (1.02–1.10) | 1.15 (1.11–1.20) | 1.34 (1.29–1.40) |
| Covariate + IDPs | 1.07 (1.01–1.13) | 1 (ref) | 1.09 (0.91–1.29) | 1.14 (0.96–1.36) | 1.17 (0.98–1.39) | 1.29 (1.09–1.53) |
| **High-sensitivity C-reactive protein** | | | | | | |
| Covariate-adjusted | 1.31 (1.30–1.32) | 1 (ref) | 1.03 (1.00–1.07) | 1.14 (1.10–1.17) | 1.39 (1.34–1.43) | 1.96 (1.90–2.02) |
| Covariate + lipid profile | 1.30 (1.29–1.32) | 1 (ref) | 1.03 (0.99–1.07) | 1.13 (1.09–1.72) | 1.39 (1.34–1.44) | 1.92 (1.86–1.99) |
| Covariate + IDPs | 1.17 (1.11–1.24) | 1 (ref) | 1.08 (0.89–1.30) | 1.25 (1.04–1.50) | 1.38 (1.15–1.65) | 1.50 (1.25–1.80) |

**Supplementary Table 10. Hazard ratios for all-cause death according to standard deviation increase and quintiles of increasing baseline inflammatory biomarker levels.** Glycoprotein acetyls and C-reactive protein were log-transformed and scaled. Lipid profile included: total cholesterol, LDL, VLDL, HDL, remnant cholesterol, total triglycerides, Lp(a). Imaging-derived phenotypes (IDPs) included: indexed left ventricle (LV) end-systolic volume, indexed LV mass, LV global wall thickness, right ventricle (RV) end-diastolic volume, RV end-systolic volume, RV ejection fraction (EF), indexed left atrium minimum volume, left atrium EF, indexed right atrium stroke volume, ascending aorta minimum area, descending aorta distensibility, global circumferential strain, circumferential peak-diastolic strain rate, longitudinal peak-diastolic strain rate. All models were adjusted for body surface area, sex, age, age$^2$, age:sex interaction, statin use. Participants were followed from enrollment until first occurrence of major cardiovascular events (MACE) for the lipid-adjusted model, or from the date of the cardiac magnetic resonance imaging until first MACE for the IDPs-adjusted model.

### Supplementary figures

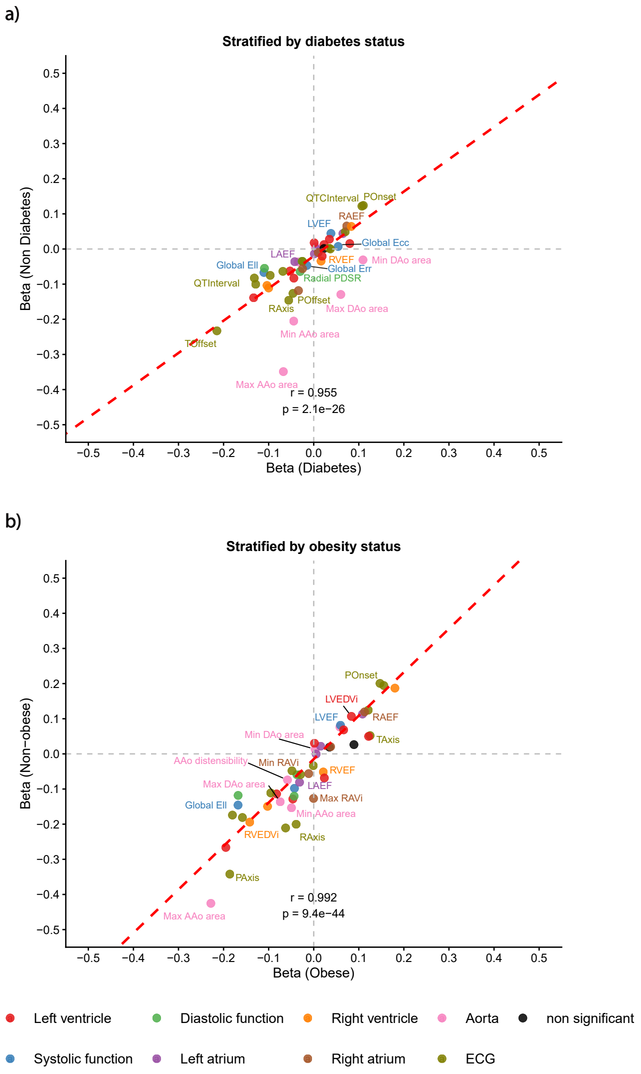


**Supplementary Figure 1. Correlation plot of multivariable linear regression coefficients for the effect of GlycA on IDPs and ECG variables stratified by diabetes (a) and obesity (b) status.** Horizontal and vertical jitters are added to reduce points overlap. The 20 variables with the highest residuals are labeled. **Abbreviations:** AAo, ascending aorta; DAo, descending aorta; ECG, electrocardiogram; EDVi, end-diastolic volume indexed; Ell, longitudinal strain; Ecc, circumferential strain; Err, radial strain; ESVi, end-systolic volume indexed; GlycA, glycoprotein acetyl; IDP, imaging-derived phenotype; LA, left atrium; LAEF, left atrial ejection fraction; LASVi, left atrial stroke volume indexed; LAVmax, maximum left atrial volume indexed; LAVmin, minimum left atrial volume indexed; LV, left ventricle; LVCO, left ventricular cardiac output; LVCi, left ventricular cardiac index; LVEDVi, left ventricular end-diastolic volume indexed; LVEF, left ventricular ejection fraction; LVESVi, left ventricular end-systolic volume indexed; LVMi, left ventricular mass indexed; LVSVi, left ventricular stroke volume indexed; MAP, mean arterial pressure; PDSR, peak diastolic strain rate; P, P wave; Q, Q wave; QRS, QRS interval; R, R wave; RA, right atrium; RAEF, right atrial ejection fraction; RASVi, right atrial stroke volume indexed; RAVmax, maximum right atrial volume indexed; RAVmin, minimum right atrial volume indexed; RV, right ventricle; RVEDVi, right ventricular end-diastolic volume indexed; RVEF, right ventricular ejection fraction; RVESVi, right ventricular end-systolic volume indexed; RVSVi, right ventricular stroke volume indexed; SBP, systolic blood pressure; SVi, stroke volume indexed; T, T wave; WT, myocardial wall thickness.

a)

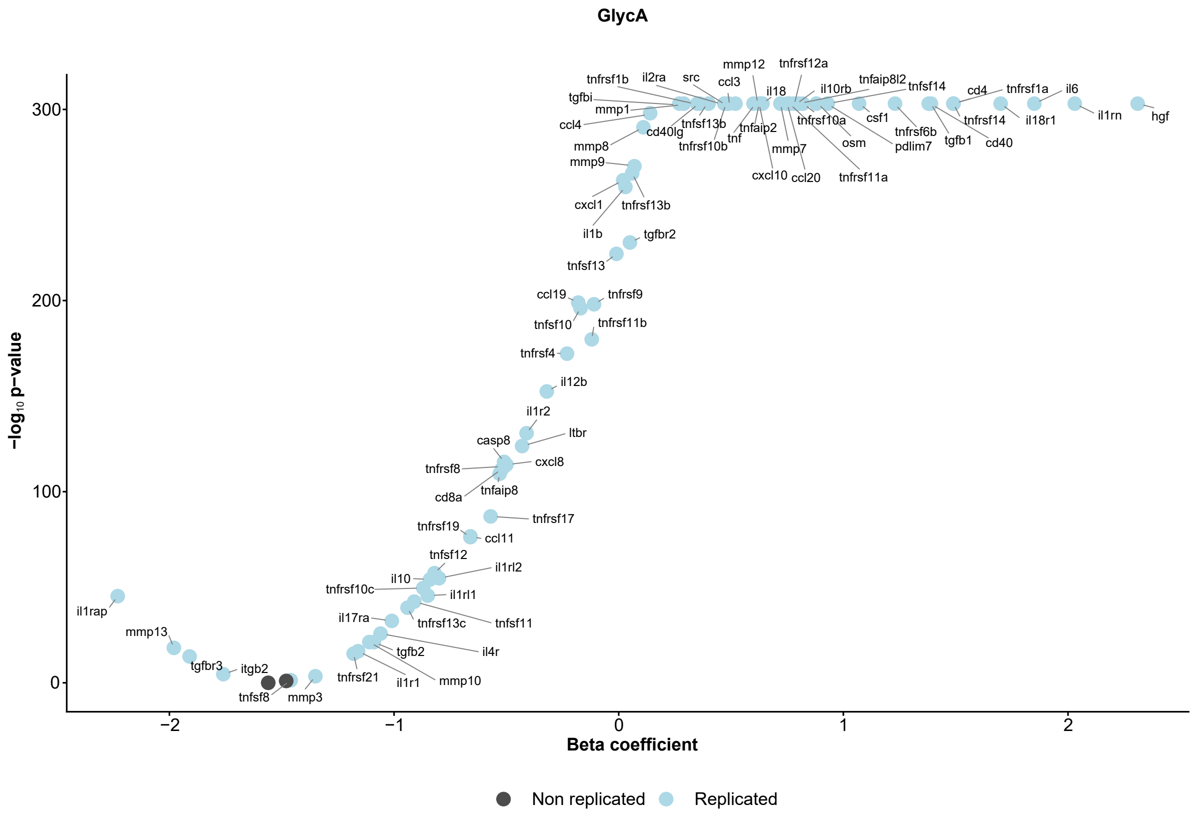


b)

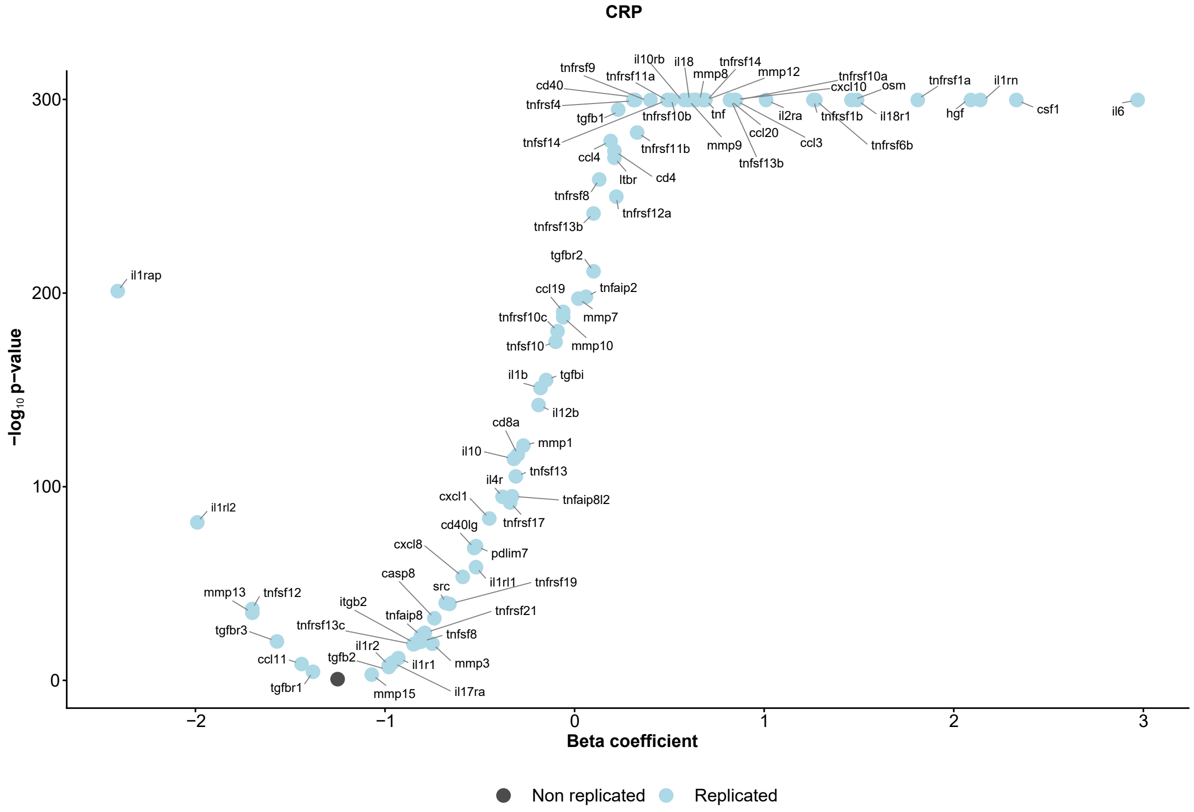


**Supplementary Figure 2. Relationship between aspecific inflammatory biomarkers and inflammatory proteins**. **a)** Multivariable linear regression coefficients for the effect of each protein on GlycA levels. Each model was built considering GlycA as the outcome variable and each protein (separately) as predictors, and adjusted for BSA, age, sex, age:sex interaction, age$^2$, and diabetes status. $\beta$ coefficients are scaled. Black dots correspond to FDR-corrected p-value < 0.05. **b)** Multivariable linear regression coefficients for the effect of each protein on CRP levels. Each model was built considering CRP as the outcome variable and each protein (separately) as predictors, and adjusted for BSA, age, sex, age:sex interaction, age$^2$, and diabetes status. $\beta$ coefficients are scaled. Black dots correspond to FDR-corrected p-value < 0.05. **Abbreviation:** BSA, body mass index; CRP, C-reactive protein; GlycA, glycoprotein acetyl.

a)

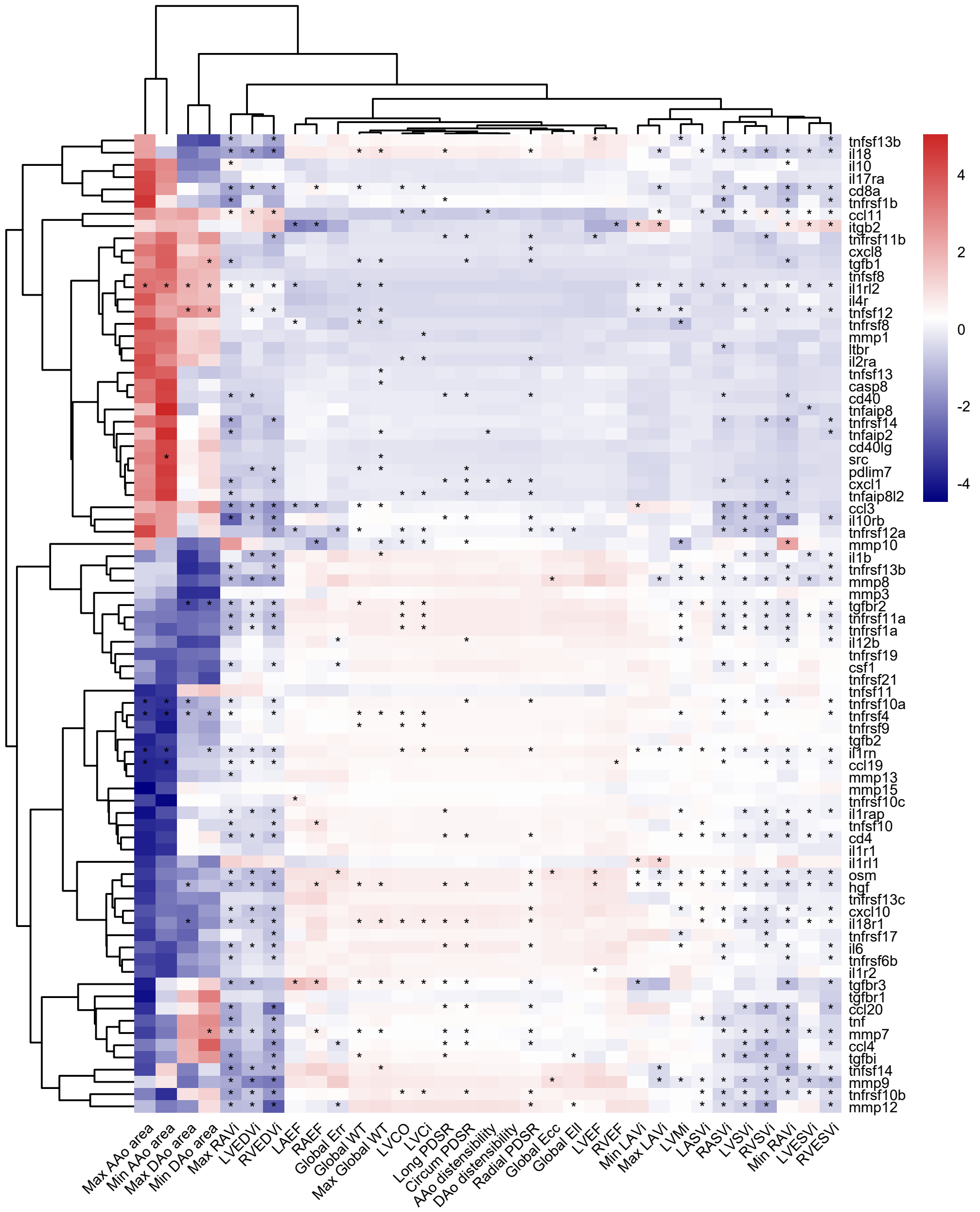
tnfsf13b
il18
il10
il17ra
cd8a
tnfrsf1b
ccl11
itgb2
tnfrsf11b
cxcl8
tgfb1
tnfsf8
il1rl2
il4r
tnfsf12
tnfrsf8
mmp1
ltbr
il2ra
tnfsf13
casp8
cd40
tnfaip8
tnfrsf14
tnfaip2
cd40lg
src
pdlim7
cxcl1
tnfaip8l2
ccl3
il10rb
tnfrsf12a
mmp10
il1b
tnfrsf13b
mmp8
mmp3
tgfbr2
tnfrsf11a
tnfrsf1a
il12b
tnfrsf19
csf1
tnfrsf21
tnfsf11
tnfrsf10a
tnfrsf4
tnfrsf9
tgfb2
il1rn
ccl19
mmp13
mmp15
tnfrsf10c
il1rap
tnfsf10
cd4
il1r1
il1rl1
osm
hgf
tnfrsf13c
cxcl10
il18r1
tnfrsf17
il6
tnfrsf6b
il1r2
tgfbr3
tgfbr1
ccl20
tnf
mmp7
ccl4
tgfbi
tnfsf14
mmp9
tnfrsf10b
mmp12
4
2
0
-2
-4
Max AAo area
Min AAo area
Max DAo area
Min DAo area
Max RAVi
LVEDVi
RVEDVi
LAEF
RAEF
Global Err
Global WT
Max Global WT
LVCO
LVCi
Long PDSR
Circum PDSR
AAo distensibility
DAo distensibility
Radial PDSR
Global Ecc
Global Ell
LVEF
RVEF
Min LAVi
Max LAVi
LVMi
LASVi
RASVi
LVSVi
RVSVi
Min RAVi
LVESVi
RVESVi

b)

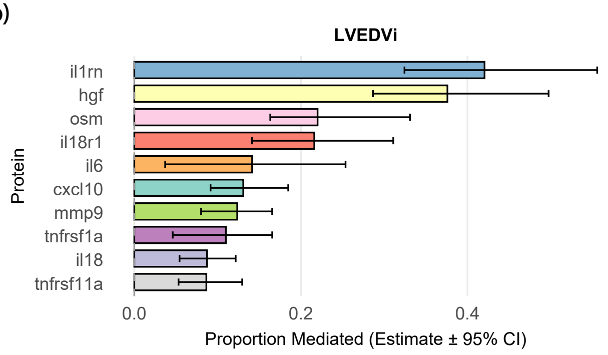
LVEDVi
il1rn
hgf
osm
il18r1
il6
cxcl10
mmp9
tnfrsf1a
il18
tnfrsf11a
Protein
0.0
0.2
0.4
Proportion Mediated (Estimate ± 95% CI)

c)

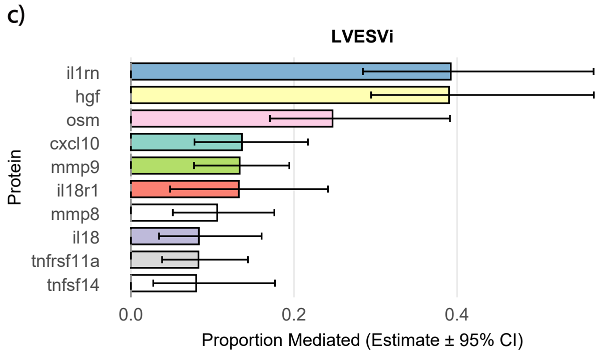
LVESVi
il1rn
hgf
osm
cxcl10
mmp9
il18r1
mmp8
il18
tnfrsf11a
tnfsf14
Protein
0.0
0.2
0.4
Proportion Mediated (Estimate ± 95% CI)

d)

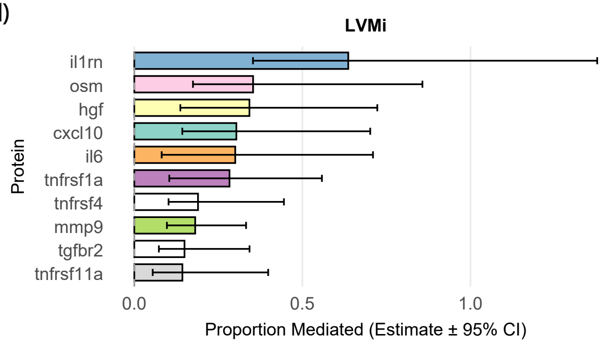
LVMi
il1rn
osm
hgf
cxcl10
il6
tnfrsf1a
tnfrsf4
mmp9
tgfbr2
tnfrsf11a
Protein
0.0
0.5
1.0
Proportion Mediated (Estimate ± 95% CI)

e)

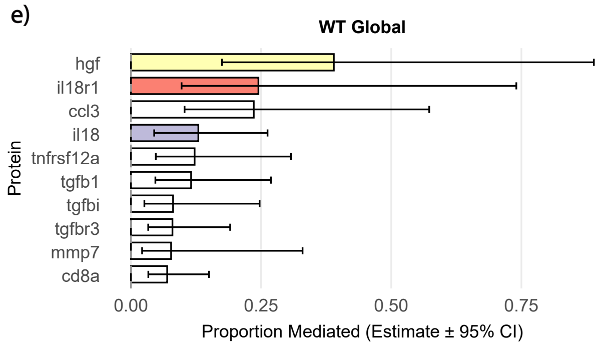
WT Global
hgf
il18r1
ccl3
il18
tnfrsf12a
tgfb1
tgfbi
tgfbr3
mmp7
cd8a
Protein
0.00
0.25
0.50
0.75
Proportion Mediated (Estimate ± 95% CI)

**Supplementary Figure 3. Proteins mediating the effect of hsCRP on cardiac geometry**. **a)** Heatmap showing the scaled coefficients for the average causal mediation effect of each protein on each IDP. Each model included log-transformed and scaled CRP as the exposure, one protein as the mediator, and one IDP as the outcome. Models were adjusted for BSA, sex, age, age$^2$, the age−sex interaction, and prevalent diabetes. (*) denotes an FDR-adjusted p-value < 0.05. The bottom panel reports the proportion of the mediated effect for the top 10 proteins for LVEDVi (**b**), LVESVi (**c**), LVMi (**d**), and global WT (**e**). **Abbreviations:** AAo, ascending aorta; BSA, body surface area; hsCRP, high-sensitivity C-reactive protein; DAo, descending aorta; EDVi, end-diastolic volume indexed; Ell, longitudinal strain; Ecc, circumferential strain; Err, radial strain; ESVi, end-systolic volume indexed; GlycA, glycoprotein acetyl; IDP, imaging-derived phenotype; LA, left atrium; LAEF, left atrial ejection fraction; LASVi, left atrial stroke volume indexed; LAVmax, maximum left atrial volume indexed; LAVmin, minimum left atrial volume indexed; LV, left ventricle; LVCO, left ventricular cardiac output; LVCi, left ventricular cardiac index; LVEDVi, left ventricular end-diastolic volume indexed; LVEF, left ventricular ejection fraction; LVESVi, left ventricular end-systolic volume indexed; LVMi, left ventricular mass indexed; LVSVi, left ventricular stroke volume indexed; MAP, mean arterial pressure; PDSR, peak diastolic strain rate; RA, right atrium; RAEF, right atrial ejection fraction; RASVi, right atrial stroke volume indexed; RAVmax, maximum right atrial volume indexed; RAVmin, minimum right atrial volume indexed; RV, right ventricle; RVEDVi, right ventricular end-diastolic volume indexed; RVEF, right ventricular ejection fraction; RVESVi, right ventricular end-systolic volume indexed; RVSVi, right ventricular stroke volume indexed; SVi, stroke volume indexed; WT, myocardial wall thickness.

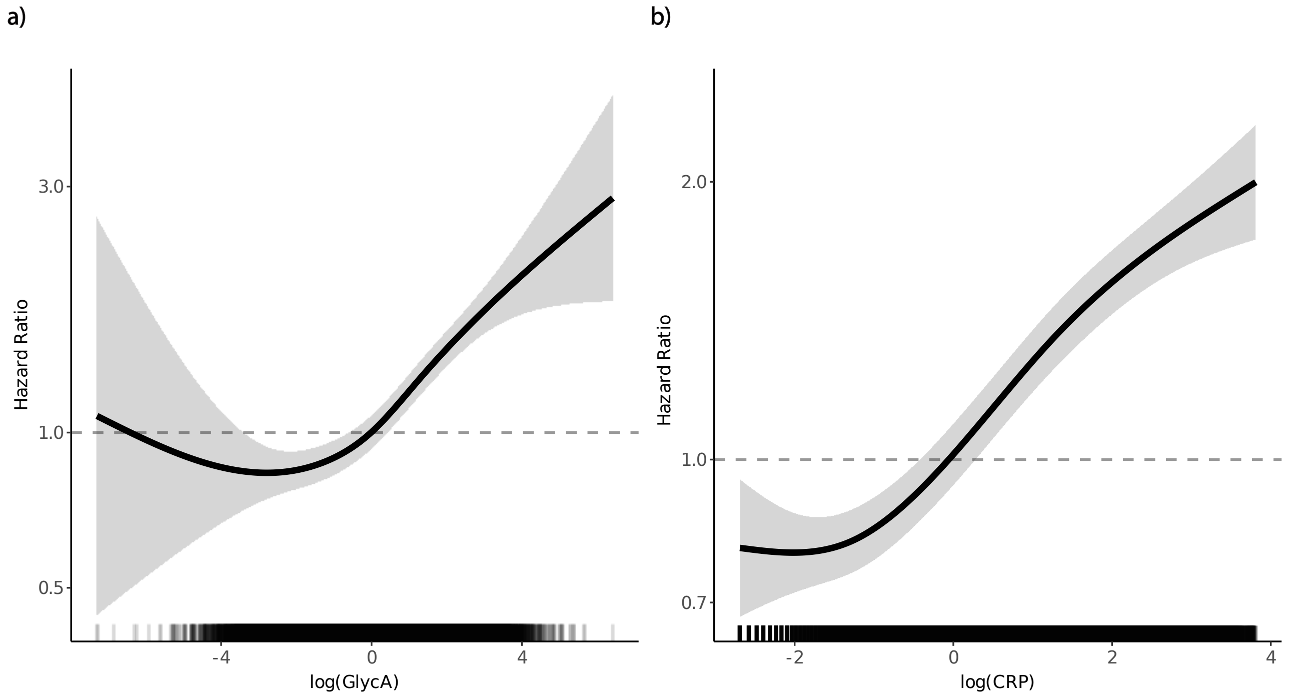


**Supplementary Figure 4. Spline curves for MACE according to changes in inflammatory biomarkers.** The curves represent the hazard ratios for MACE, with overlaid 95% confidence intervals, across the range of standard deviation changes in log-transformed and scaled GlycA **(A)** and hsCRP **(B)**, considering a median follow-up of 15.3 years. **Abbreviations:** hsCRP, high-sensitivity C-reactive protein; GlycA, glycoprotein acetyls; MACE, major adverse cardiovascular events.

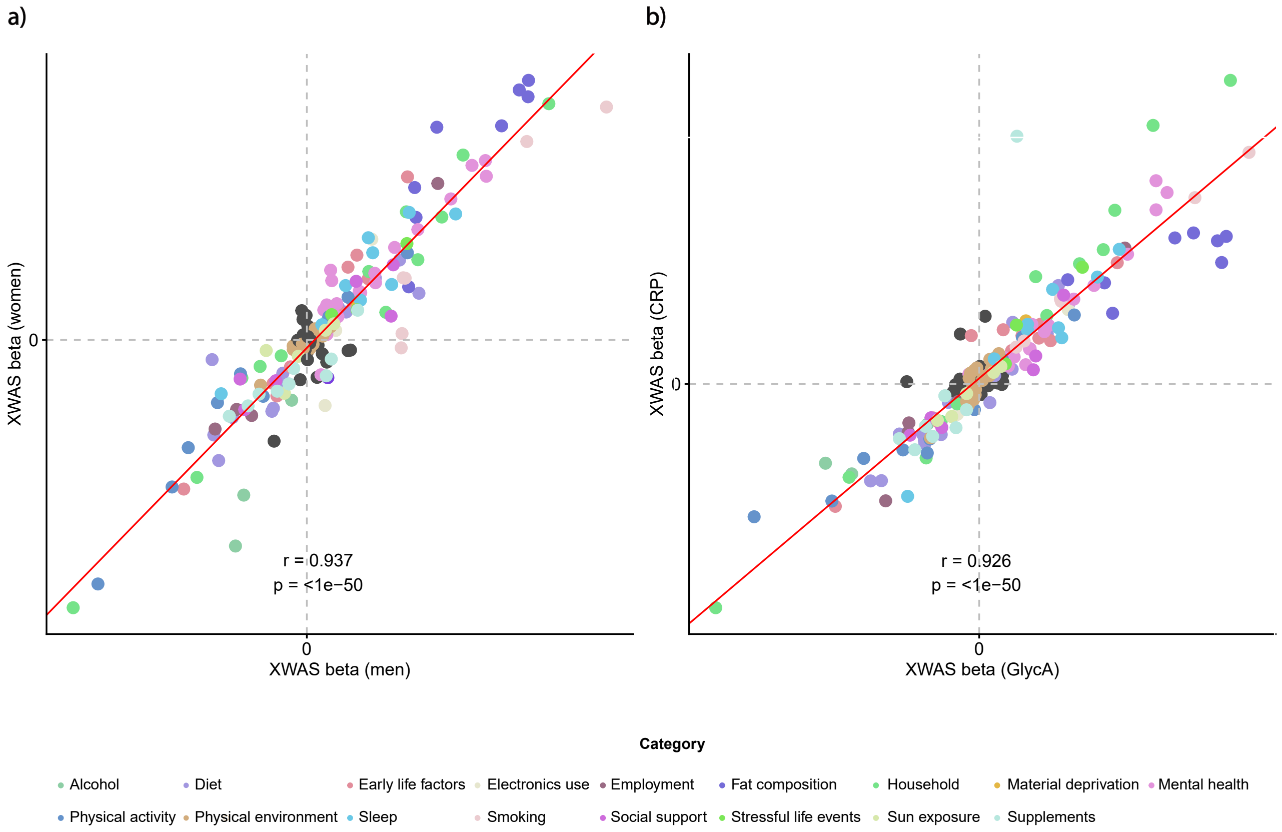


**Supplementary Figure 5. Correlation plot of XWAS coefficients for the effects of exposures on inflammatory biomarkers. a)** Stratified by sex. **b)**, Using hsCRP instead of GlycA. **Abbreviations:** CRP, high-sensitivity C-reactive protein; GlycA, glycoprotein acetyl; XWAS, exposome-wide association study

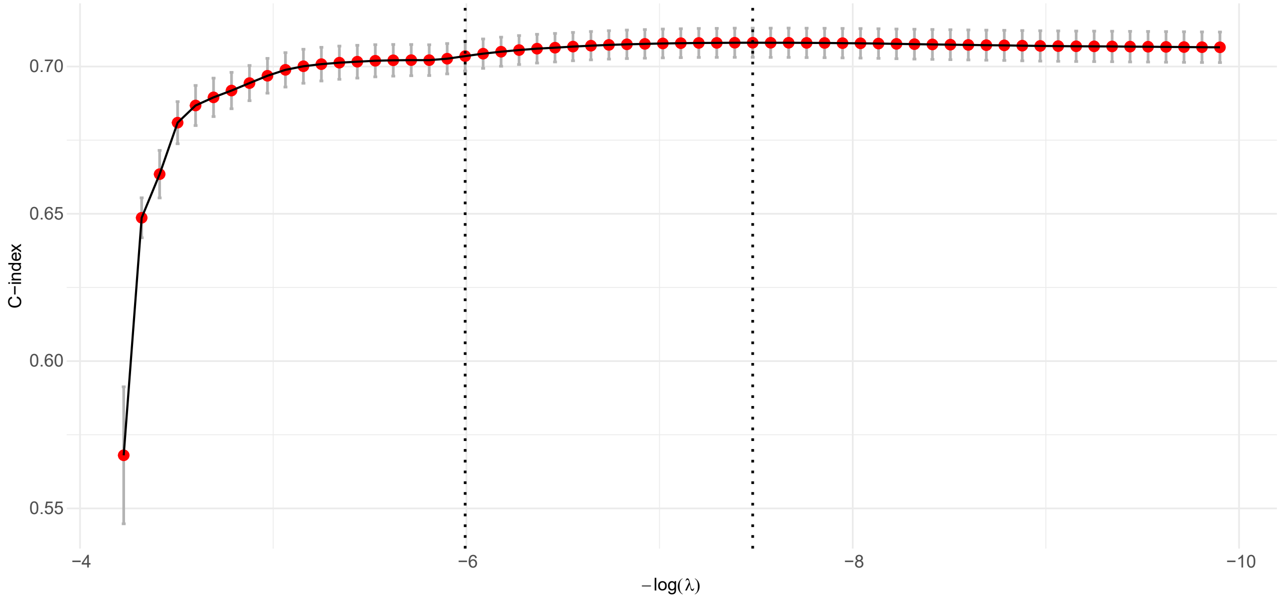


**Supplementary Figure 6. Feature selection by least absolute shrinkage and selection operator.** Feature selection was performed using 10-fold cross-validation to identify the best regularization parameter $\lambda$ defined by the $\lambda$ giving the highest AUC−1 standard error. Using the best $log(\lambda)$ parameter of $5.63\times10^{-4}$, 14 variables were selected.
